# Impact of newborn screening on survival and neurocognitive outcome in classic isovaleric aciduria: a meta-analysis

**DOI:** 10.1101/2025.04.09.25325525

**Authors:** Anna T. Reischl-Hajiabadi, Sven F. Garbade, Florian Gleich, Elena Schnabel-Besson, Roland Posset, Matthias Zielonka, Georg F. Hoffmann, Stefan Kölker, Ulrike Mütze

## Abstract

**Purpose:** Classic isovaleric aciduria (cIVA) is a rare inherited metabolic disorder characterized by recurrent life-threatening metabolic decompensations and neurocognitive impairment in untreated patients. This meta-analysis aims to assess the impact of early diagnosis on mortality and neurocognitive outcome.

**Methods:** A systematic literature search for articles published until 2022 was conducted following PRISMA protocol guidelines. We investigated effects on clinical outcome and survival, analyzing outcome parameters using meta-analytical measures and estimating effect sizes with a random-effects model.

**Results:** Overall, 20 studies were included, reporting on 240 individuals with cIVA. Individuals identified by NBS presented with a lower frequency of neurological symptoms (13.0% versus 44.9%; *P* = 0.0040) and developmental delay (6.1% versus 51.2%; *P* < 0.0001), and had a lower mortality rate (1.1% versus 10.9%; *P* = 0.0320). The quality of national healthcare systems did not have a measurable impact on neurocognitive outcome and mortality. Despite the beneficial effect of NBS on clinical outcome and mortality, it could not reliably prevent the manifestation of neonatal decompensation in all individuals with cIVA identified by NBS.

**Conclusions:** Early diagnosis through NBS is essential for the timely initiation of therapy and for improving outcomes and survival rates in individuals with cIVA.

## Introduction

Isovaleric aciduria (IVA, OMIM #243500) is a rare inborn error of leucine metabolism due to bi-allelic pathogenic variants in the *IVD* gene (cytogenetic location: 15q15.1), resulting in inherited deficiency of the mitochondrial enzyme isovaleryl-CoA dehydrogenase (EC 1.3.99.10) and, subsequently, in mitochondrial accumulation of toxic isovaleryl-CoA and other metabolites. Untreated individuals with classic IVA (cIVA) often present with life-threatening acute metabolic decompensations in the first days of life, resulting in high neonatal mortality (1, 2). These episodes are precipitated by fasting, infectious disease, or increased protein intake also after the newborn period (3) and lead to impaired neurocognitive development in infants surviving the initial decompensation (1, 4). Treatment consists of a low protein diet with or without leucine-free amino acid supplementation, detoxification of isovaleryl-CoA through application of carnitine or glycine, and avoidance of catabolic episodes (4, 5).

Aiming to improve neonatal survival and neurocognitive outcome through early diagnosis and pre-symptomatic start of treatment, IVA was introduced to newborn screening (NBS) programs more than 20 years ago. NBS not only identified individuals with clinically severe cIVA but also with an attenuated disease variant (aIVA), which had been unknown in the pre-screening era (4, 6) and was mainly associated with a common *IVD* gene variant (c.932C>T; Transcript NM_002225.5) (4). Consequently, the estimated birth prevalence for IVA increased from 1 in 280,000 in pre-NBS cohorts to about 1 in 100,000 newborns in NBS cohorts (4, 7, 8). While excellent outcome was shown for screened individuals with aIVA (N=70) (4), the impact of NBS for screened individuals with cIVA still remains less clear because of studies with small sample size (N=24 (4); N=10 (9); N=13 (10) N=4 (11); N=4 (12)). As far as it is known, NBS seems to reduce mortality and the overall frequency of metabolic decompensations in cIVA (1, 4, 13), while it does not seem to reliably protect against neonatal metabolic decompensations.

To better understand the impact of NBS on mortality and neurocognitive outcome in individuals with cIVA, the aim of this meta-analysis is the systematic evaluation of reported screened and unscreened cohorts of individuals with cIVA.

## Materials and Methods

### Search strategy and study design

A systematic literature search was conducted according to the PRISMA 2020 guidelines (14) (Prisma 2020 flow; Figure 1), exploring PubMed, Web of Science, and Cochrane Library for reports published since the first description of IVA patients by Tanaka and colleagues in 1966 (15) until December 31^st^, 2022. The search utilized the MeSH terms “isovaleric acidemia” and “isovaleric aciduria”. For the reviewing process, the PRISMA 2020 flow chart and protocol (14) was followed which was finalized through manual searches and reaching out to authors (Figure 1).

**Figure 1:**
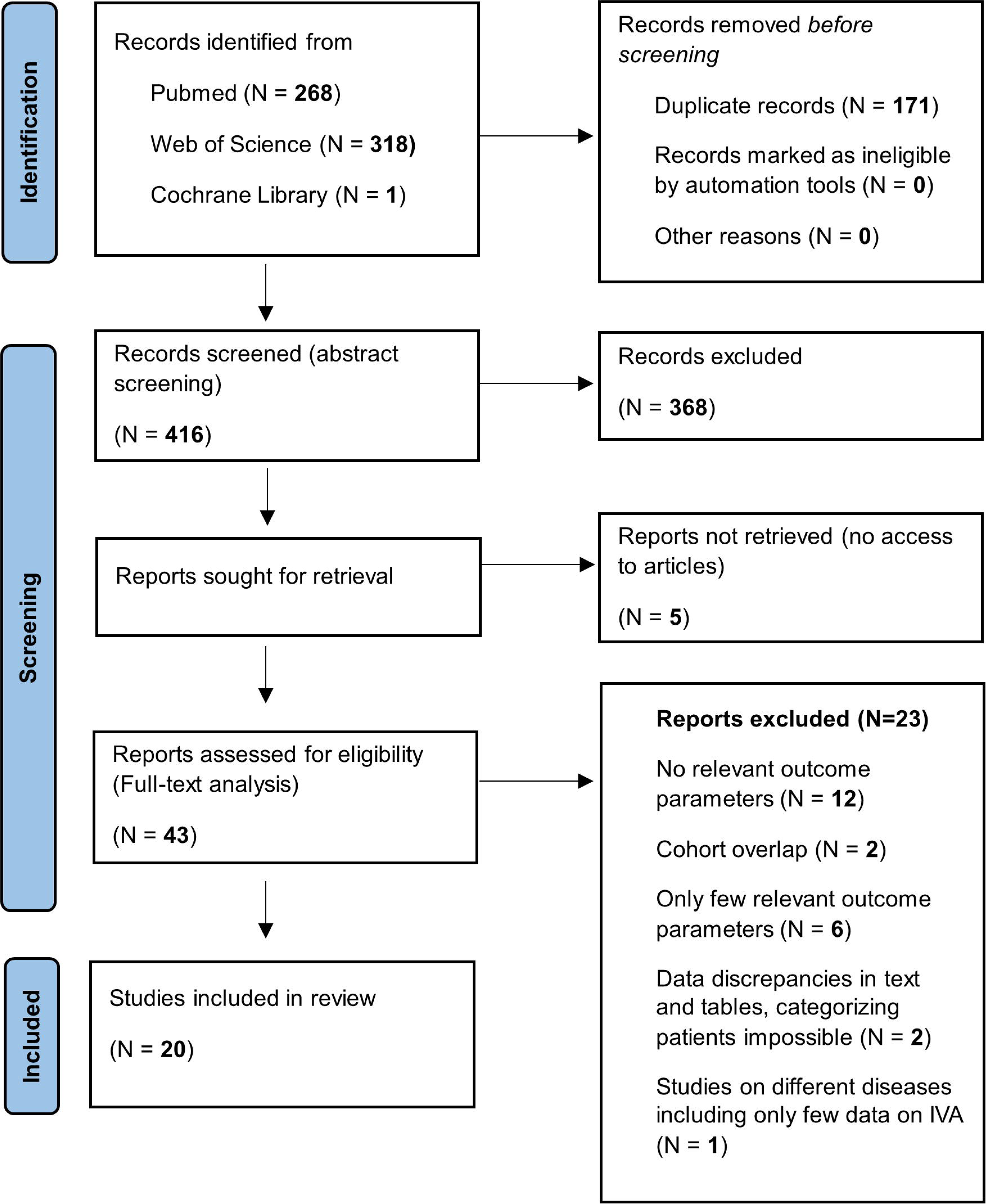
PRISMA flow diagram demonstrating the flow of information of the meta-analysis. Finally, a total of 20 studies were included in the analyses. 9 studies included data on individuals with cIVA identified by NBS (cIVA-NBS) and 17 studies included data on individuals diagnosed after onset of symptoms (cIVA-Sympt).

### Inclusion and exclusion criteria

Prior to inclusion in the study analysis, the abstracts of all identified publications were screened by at least two reviewers independently regarding the following aspects: (1) report of individuals with confirmed diagnosis of IVA, (2) presentation of clinical outcome indicators for each patient. In analogy, abstracts selected to be eligible for the analysis were checked in full text by at least two independent reviewers according to the following criteria: (A) cohort size of at least three individuals with IVA, (B) report of variables according to the data model of this study (e.g. mortality, metabolic decompensation, age, data on NBS, neurological signs and symptoms, cognitive development, IQ values), (C) minimal set of core variables (e.g. diagnostic mode, age at diagnosis, age at the last follow-up, metabolic decompensation, data on cognitive or clinical development, mortality) extractable for analysis. For multiple publications on the same study cohort, the most comprehensive or most recent publication was included in the analysis. The reasons for excluding studies from the analysis are specified in Supplementary Table 1.

Detailed evaluation on the risk of bias for each study is summarized in Supplementary Figure 1 in analogy to previous studies (16–18).

### Data model

Data was extracted from the manuscript’s text, tables, and figures by full-text analysis. The data model included the following parameters/variables: sex, disease variant (cIVA or aIVA; definition used see below), age at diagnosis, age at the last follow-up, mode of diagnosis (via NBS [including high-risk (HR) family screening in an asymptomatic stage within the first three months of life] or following the onset of symptoms), data on clinical phenotype (e.g. age at onset of symptoms or metabolic decompensation, isovaleryl-carnitine (C5) at NBS/diagnosis, maximum plasma ammonia level (NH_3_) at first metabolic decompensation, age at start of treatment, neurological symptoms at last follow-up, age of death if applicable) and cognitive parameters (results of IQ-based tests and Denver Developmental Screening Test [DDST], and presence of developmental delay at last follow-up).

Data were extracted for subgroups defined by mode of diagnosis (NBS or diagnosis after onset of symptoms) and confirmed disease, the presence of symptoms at diagnosis (symptomatic or asymptomatic), and the disease severity (cIVA or aIVA) (Supplementary Table 2). Since data quality and the number of reported variables varied among studies, not all selected studies could be included for every analysis. If a specific variable was not reported in a study, it was marked as “not reported” (NR).

As the included reports on individuals with IVA have been published over a long period and in different countries, we aimed to investigate whether differences in national healthcare systems had an impact on the clinical and neurocognitive outcome. The Healthcare Access and Quality (HAQ) Index is a metric introduced by the Global Burden of Diseases, Injuries, and Risk Factors Study (19–22). It considers indicators such as mortality rates, healthcare services, disease control, quality of care, access to medications, infrastructure and healthcare financing, and was used to rank the selected articles included in this meta-analysis according to the country where data collection was performed and according to the study period (Supplementary Table 2). The HAQ Index comprises values from 0 (poor) to 100 (excellent) and has been released approximately every five years since 1990 for all countries worldwide (19–22).

### Classification

Screened individuals with IVA were classified as aIVA if at least one of the following criteria was met, as previously described (4): homozygosity or compound heterozygosity for the *IVD* gene variant c.932C>T (transcript NM_002225.5), C5 <6 μmol/L in the first NBS sample and absence of neonatal metabolic decompensation. Subsequently, data sets of individauls with aIVA were excluded from further analysis.

Individuals who exhibited IVA-related symptoms or experienced at least one acute metabolic decompensation until the last follow-up visit were classified as “symptomatic”, all other patients as “asymptomatic”. Metabolic decompensations and onset of other clinical symptoms were classified as early onset (EO) if they occurred in the neonatal period and as late onset (LO) if the first one occurred after this period.

### Statistical analysis

Statistical analyses were performed by using R language for statistical computation (https://www. R-project.org, Version 4.3.2). Studies with missing data were excluded from the respective analysis. The R packages ‘metafor’ version 4.4-0 (23) and ‘meta’ version 7.0-0 (24) were used for computation of meta-analysis. Proportions were logit transformed (basis e) and then a random intercept logistic regression model was applied (this is the default in package ‘meta’). For numerical outcome, a random effects model was computed. When testing subgroup differences with random effects meta regression models, we report the χ² value with degrees of freedom in parentheses followed by corresponding *P* value. In meta-regression, the slope β, z value and *P* value will be reported. As measures for heterogeneity, we report Cochran’s *Q* (in parentheses degrees of freedom) followed by corresponding *P* value and *I^2^*: Cochran’s *Q* given the alternate hypothesis is of heterogeneity, and *I^2^*, which estimates the percentage of variation across studies that is due to heterogeneity rather than chance. For publication bias, the Egger and Begg tests were calculated. If the number of studies was less than 10, no test was performed. Results were displayed as forest and funnel plots. *P* values <0.05 were considered as statistically significant. The results are presented as follows: In *Description of included studies*, the selected studies are outlined using descriptive measures without applying a meta-analytic integration method. In the *Meta-analysis* section, effect sizes are estimated using a random effects model.

## Results

### Study sample

Out of 416 literature records that were identified and screened, 5 reports could not be retrieved and 368 records were excluded following abstract assessment. Subsequently, 43 articles underwent full-text evaluation for eligibility, and 23 of them were excluded for various specific reasons (Supplementary Table 1; Figure 1). Ultimately, 20 publications from 23 countries (1, 2, 4, 9–12, 25–37), reporting on individuals with cIVA identified either by NBS (cIVA-NBS, N=60; 9 publications (4, 9–12, 25, 28, 31, 34)) or following the onset of symptoms (cIVA-Sympt; N=180; 17 publications (1, 2, 10–12, 26–37)), were included in the quantitative synthesis (Figure 1; Supplementary Table 2), overall, 240 patients with cIVA. Since a multi-national study on cIVA from 14 countries (10) showed partial overlap with a national observational study (4), raw data was reviewed and duplicates were deleted. Data on 91 individuals with aIVA were excluded for further analysis. Detailed information on the study population is displayed in Supplementary Table 2.

### Age at diagnosis and disease onset

#### Description of included studies

In the cIVA-NBS group (N=60; 39 female, 21 male), individuals were diagnosed at a weighted mean age of 10 days (range 4-27 days) and were followed for a weighted mean duration of 95 months (range 0.4-129 months). Thirty-seven percent (N=22) of them were already symptomatic at first NBS report or confirmation of suspected diagnosis. Overall, 62% (N=37) of the cIVA-NBS group presented with at least one metabolic decompensation at a weighted mean age of 150 days (range 8-175 days) (EO group: weighted mean age of 5 days; LO group: weighted mean age of 731 days) or disease-specific symptoms, the majority of them (70%; N=26) already neonatally. Thirty-eight percent of the cIVA-NBS group (N=23) remained asymptomatic until the last follow-up. In the cIVA-Sympt group (N=180, 83 female, 93 male, 4 sex not reported), the weighted mean age at the last follow-up was 128 months (range 22-258 months). In this group, 117 patients (65%) presented with an EO and 63 (35%) with a LO of symptoms. The first metabolic decompensation occurred at a weighted mean age of 375 days (range 5-893 days) (EO: weighted mean age of 7 days; LO: weighted mean age of 998 days) and diagnosis was confirmed at a weighted mean age of 727 days (range 18-1340 days) (EO: weighted mean age of 353 days; LO: weighted mean age of 1313 days).

#### Meta-analysis

A comparison of the diagnosis groups showed that individuals of the cIVA-NBS group were diagnosed at a younger age (log mean 2.26, back-transformed 9.58 days; 95% log confidence interval (95%-CI): 1.54-2.98) compared to those of the cIVA-Sympt group (log mean 6.10, back-transformed 445.9 days; 95%-log CI 5.47-6.74; [χ²(1) = 61.3, *P* < 0.0001; Q(18) = 26,969.3, *P* = 0; I^2^ = 99.9%]). Test on publication bias found conflicting results: Egger’s test detects publication bias (t(17) = 2.29, *P* = 0.035), but Begg’s test does not (z = −1.29, *P* = 0.196). So, a publication bias is likely for this analysis. Further, individuals of the cIVA-NBS group were less often symptomatic at diagnosis (cIVA-NBS: 31%; 95%-CI 0.11-0.51; cIVA-Sympt: 99.8%; 95%-CI 0.97-1.0; [χ²(1) = 43.62, *P* < 0.0001; Q(23) = 129.68, *P* < 0.0001; I^2^ = 82.3%]). Publication bias was present (Egger’s test: t(22) = −2.96, *P* = 0.007; Begg’s test: z = −4.43, *P* < 0.0001). Evaluation of the age at first metabolic decompensation, however, did not show differences between the modes of identification (cIVA-NBS: log mean 3.59, back-transformed 36.2 days; 95%-CI 0.54-6.66; cIVA-Sympt: log mean 4.89, back-transformed 132.9 days; 95%-CI 3.18-6.61; [χ²(1) = 0.52, *P* = 0.4687; Q(8) = 1,966.08, p = 0; I^2^ = 99.6%]).

### Survival and neurocognitive outcome

#### Description of included studies

In analogy to the age at diagnosis, the weighted mean age at treatment start was 8 days (range 1-27 days) in the cIVA-NBS group compared to 582 days (range 244-1257 days) in the cIVA-Sympt group.

#### Meta-analysis

This demonstrates that NBS enables a much earlier start of treatment (log mean 2.47, back-transformed 11.82 days; 95%-CI 0.90-4.04) compared to unscreened populations (log mean 6.44, back-transformed 626.4 days; 95%-CI 5.94-6.93; [χ²(1) = 22.24, *P* < 0.0001; Q(7) = 14,862.88, *P* = 0; I^2^ = 100%]).

#### Description of included studies

Overall, 29 of 180 children of the cIVA-Sympt group died, resulting in a biased and uncorrected mortality rate of 16.1%, and for cIVA-NBS group 2 of 60 children (3.3%).

#### Meta-analysis

To obtain a more accurate estimate of the mortality rates, a random effects model was used. Based on this model, an adjusted mortality rate of 1.1% (95%-CI 0–0.06) was calculated for the cIVA-NBS group, highlighting a lower mortality rate compared to the cIVA-Sympt group (10.9%; 95%-CI 0.03–0.19; [χ²(1) = 4.6, *P* = 0.0320; Q(25) = 53.56, *P* = 0.0008; I² = 53.3%]) (Figure 2). But again, publication bias was present (Egger’s test: t(24) = 3.05, *P* = 0.0055; Begg’s test: z = 3.29, *P* = 0.0010).

**Figure 2:**
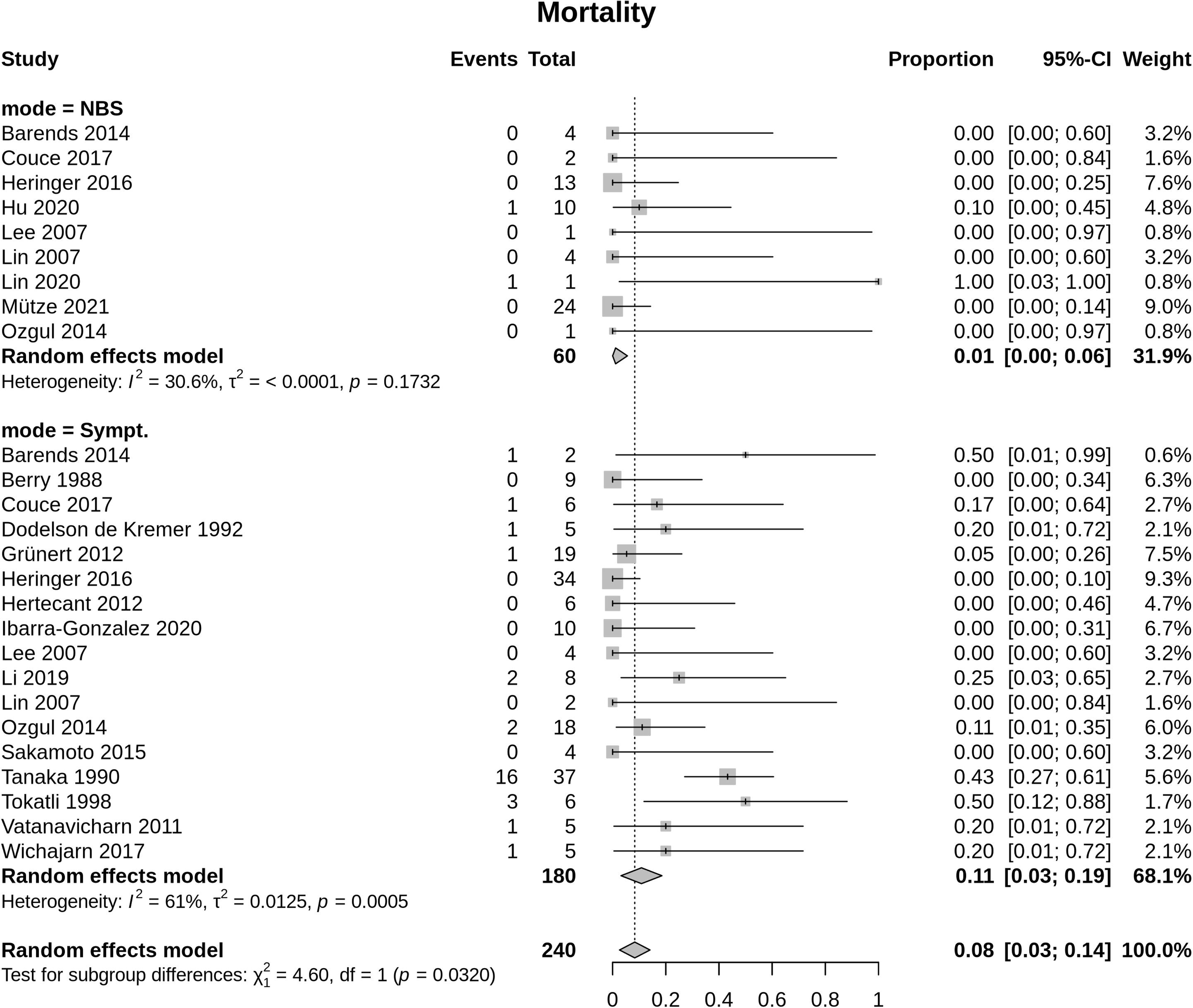
Mortality rate for classic IVA. Identification by NBS (cIVA-NBS) versus diagnosis after onset of symptoms (cIVA-Sympt). The Random effects model measures the mean weighted effect size (indicated by diamonds) and CI (confidence interval). Each square size reflects the study weight. Patients identified by NBS show a significant lower mortality rate (cIVA-NBS: 1.1%; 95%-CI 0–0.06; cIVA-Sympt: 10.9%; 95%-CI 0.03–0.19; [χ²(1) = 4.6, P = 0.0320; Q(25) = 53.56, *P* = 0.0008; I² = 53.3%]).

#### Description of included studies

Last follow-up for individuals in studies included in analysis for neurological disease manifestation was performed at a weighted mean age of 88 months in the cIVA-NBS group versus 116 months in the cIVA-Sympt group. Last follow-up for individuals in studies included in the analysis for developemental delay was done at a weighted mean age of 27 months in the cIVA-NBS group versus 118 months in the cIVA-Sympt group.

#### Meta-analysis

Evaluation of the outcome measures revealed that individuals of the cIVA-NBS group exhibited in a lower frequency of neurological disease manifestation (13.0%; 95%-CI 0-0.27) compared to the cIVA-Sympt group (44.9%; 95%-CI 0.28-0.62; [χ²(1) = 8.3, *P* = 0.0040; Q(17) = 56.78, *P* < 0.0001; I^2^ = 70.1%]; Figure 3). No publication bias was detected (Egger’s test: t(16) = 0.62, *P* = 0.545; Begg’s test: z = 0.53, *P* = 0.595). Furthermore, individuals of the cIVA-NBS group presented less often with developmental delay (cIVA-NBS: 6.1%; 95%-CI 0-0.19; cIVA-Sympt: 51.2%; 95%-CI 0.36-0.67; [χ²(1) = 18.2, *P* < 0.0001; Q(19) 80.27, *P* < 0.0001; I^2^ = 76.3%]; Figure 4). No publication bias was detected (Egger’s test: t(18) = 0.84, *P* = 0.412; Begg’s test: z = 0.52, *P* = 0.601).

**Figure 3:**
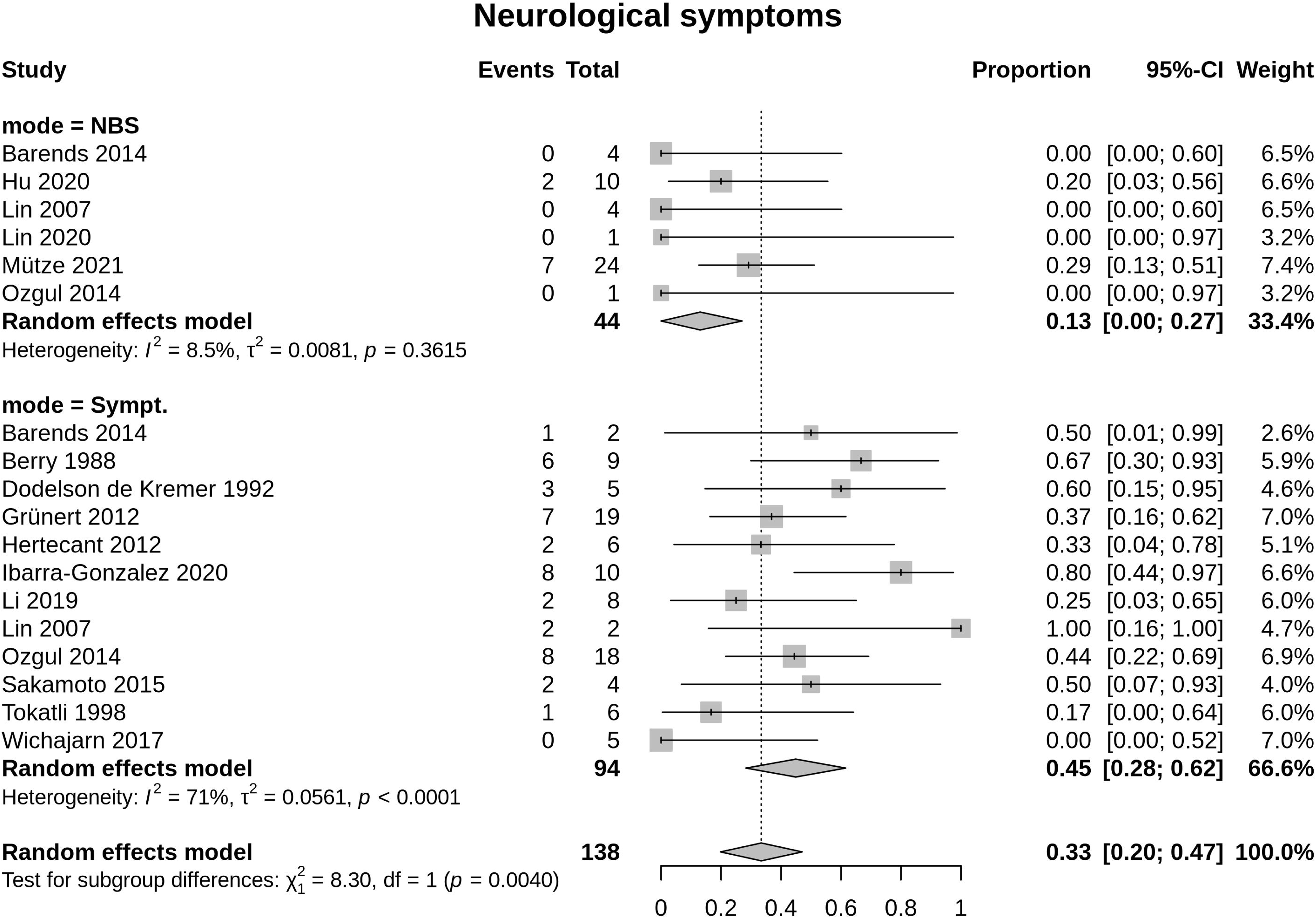
Occurrence of neurological symptoms until last follow-up in classic IVA. Identification by NBS (cIVA-NBS) versus diagnosis after onset of symptoms (cIVA-Sympt). The Random effects model measures the mean weighted effect size (indicated by diamonds) and CI (confidence interval). Each square size reflects the study weight. Patients identified by NBS show less often neurological symptoms (cIVA-NBS: 13.0%; 95%-CI 0-0.27; cIVA-Sympt group 44.9%; 95%-CI 0.28-0.62; [χ²(1) = 8.3, *P* = 0.0040; Q(17) = 56.78, *P* < 0.0001; I^2^ = 70.1%]).

**Figure 4:**
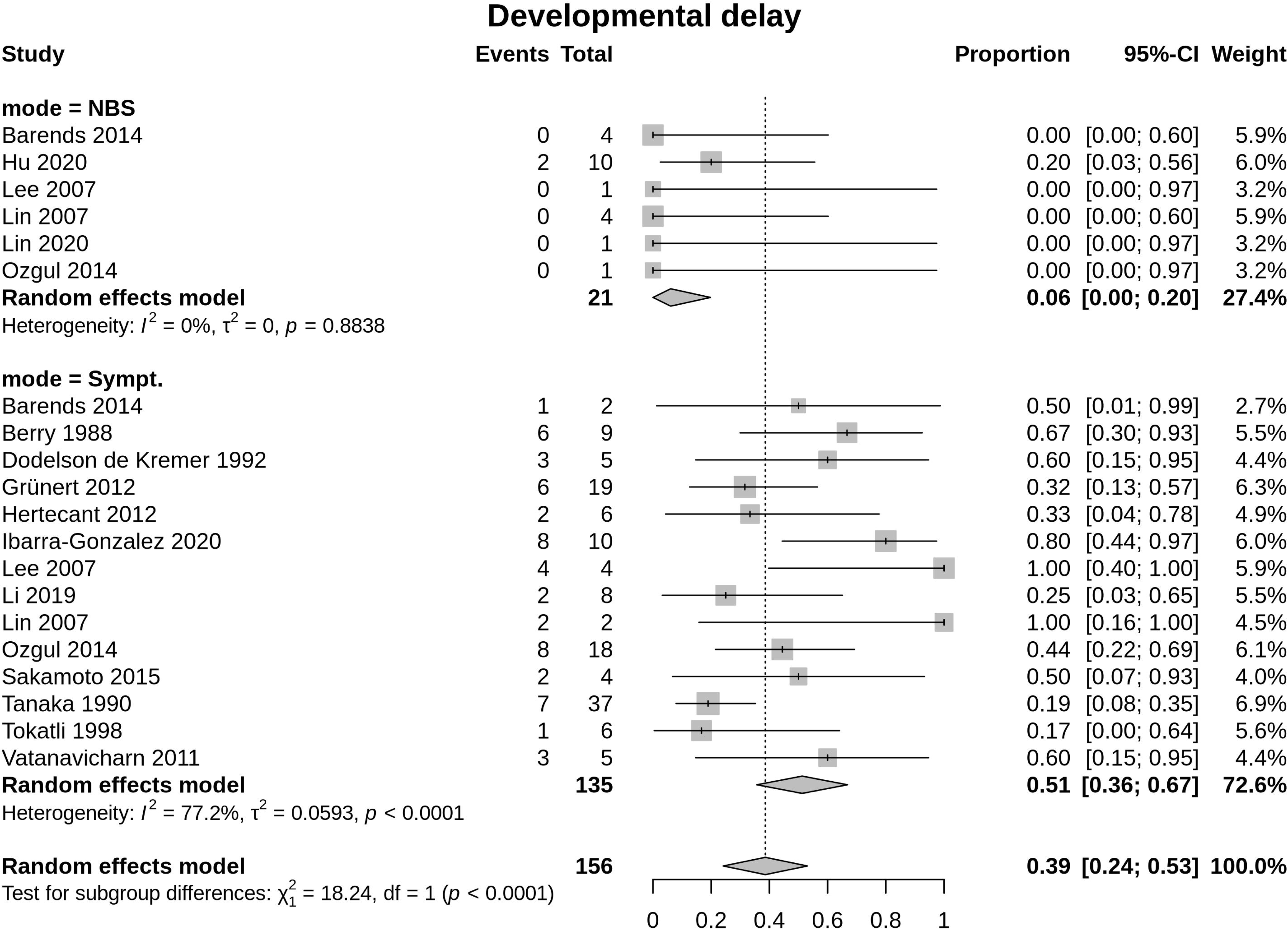
Occurrence of developmental delay until last follow-up in classic IVA. Identification by NBS (cIVA-NBS) versus diagnosis after onset of symptoms (cIVA-Sympt). The Random effects model measures the mean weighted effect size (indicated by diamonds) and CI (confidence interval). Each square size reflects the study weight. Patients identified by NBS show a significantly lower proportion of developmental delay (cIVA-NBS: 6.1%; 95%-CI 0-0.19; cIVA-Sympt: 51.2%; 95%-CI 0.36-0.67; [χ²(1) = 18.2, *P* < 0.0001; Q(19) 80.27, *P* < 0.0001; I^2^ = 76.3%].

### Health Quality Index

#### Meta analysis

Since the availability and quality of specialized services for individuals with IVA in different countries, as indicated by the Health Quality Index (HAQ) (19–22), may have an impact on diagnostic process quality, patients’ mortality, and clinical outcomes, we evaluated whether the HAQ affected the results of this study. However, the HAQ showed no measurable impact on the diagnosis modes concerning age (β_HAQ_ = -0.0783, z = -1.633, *P* = 0.1025, [Q(17) = 18,294.98, *P* = 0; I^2^ = 99.94%]) and symptoms at diagnosis (β_HAQ_ = -0.0073, z = -0.9978, *P* = 0.3184, [Q(22) = 127.75, *P* = 0; I^2^ = 93.63%]), age at treatment start (β_HAQ_ = -0.056, z = -0.8722, *P* = 0.3831, [Q(6) = 1,577.33, p = 0; I^2^ = 99.57%]), metabolic decompensations (β_HAQ_ = -0.0252, z = -0.3334, *P* = 0.7388, [Q(7) = 1,236.56, *P* = 0; I^2^ = 99.41%]), neurological symptoms (β_HAQ_ = -0.0055, z = -0.6951, *P* = 0.4870, [Q(16) = 53.25, *P* = 0; I^2^ = 71.46%]), developmental delay (β_HAQ_ = -0.0038, z = -0.4408, *P* = 6593, [Q(18) = 78.02, *P* = 0; I^2^ = 78.14%]), and mortality rate (β_HAQ_ = -0.0050, z = -1.5599, *P* = 0.1188, [Q(24) = 45.28, *P* = 0; I^2^ = 57.64%]).

## Discussion

This study on 240 individuals with cIVA from 23 countries is the most comprehensive meta-analysis on cIVA so far, spanning three decades. This study clearly demonstrates that individuals of the NBS group show reduced mortality and better clinical outcome in comparison to unscreened individuals, while it failed to demonstrate a measurable impact of the quality of national healthcare systems on the endpoints used in this study. Despite its overall benefit on survival and neurocognitive outcome, NBS does not seem to reliably protect against the manifestation of neonatal metabolic decompensations.

### NBS is the pre-requisite for improving survival and neurocognitive outcome in cIVA

This meta-analysis provides robust evidence supporting the benefit of NBS by improving survival and neurocognitive outcomes. The results of the meta-analysis confirm in a bigger cohort and at a higher evidence level the previously reported improved survival for screened indivduals with cIVA (German screened IVA cohort (4, 38)) and adds the aspect of the developmental and neurological outcome that could not be demonstrated before. Notably, these findings have to be assessed with the limitations of the divergent follow-up intervals for cIVA-NBS and cIVA-Symp cohorts with longer follow-up periods for the cIVA-Symp cohort for all endpoints (developmental delay, neurological symptoms, and mortality).

Previous studies showed that late diagnosis of cIVA in the pre-screening NBS era (1) and EO metabolic decompensations (4) were associated with cognitive impairment. In line with this, the meta-analysis demonstrates that developmental delay occurs less frequently in the NBS cohort compared to unscreened individuals with cIVA, indicating that early diagnosis and treatment through NBS reduce or mitigate EO metabolic decompensations. While previous studies with small study cohorts hinted at the benefits of early diagnosis through NBS (4) and showed that those individuals who were treated from early infancy even in the pre-screening era did not develop mental retardation (37) and more often had better normal cognitive outcome (85% early diagnosis versus 45% late diagnosis), this meta-analysis with a larger sample size confirms that NBS associated with early treatment leads to significantly improved cognitive development. However, some studies demonstrate that individuals with cIVA, particularly those who experienced severe neonatal decompensations, continue to show lower IQ values despite early diagnosis (4) but better IQ values than in the pre-screening era (1). This underscores that while NBS facilitates early treatment and reduces the risk of developmental delay, the severity of initial metabolic crises remains a determinant of long-term cognitive function.

Neurological symptoms in IVA patients have been described in smaller studies from the pre-screening era (1, 37), while a national observational study of screened patients reported favorable psychomotor development (4). For the first time, this meta-analysis directly compares both diagnostic modes and clearly demonstrates that early diagnosis and treatment significantly improve neurological outcomes. Nevertheless, the impact of severe neonatal decompensation might be a critical factor influencing neurological symptoms.

Without timely diagnosis and treatment, metabolic decompensations can escalate to lethargy, seizures, coma, and even death (39, 40). In the pre-screening era, mortality was notably high during the first metabolic decompensation (33%), with improved survival rates for those who survived the initial decompensation (3%) (1). The reduction in mortality cleary demonstrated in this meta-analysis’ results confirm that NBS significantly reduces mortality in individuals with cIVA.

### NBS mitigates but does not reliably prevent symptomatic disease courses

NBS is crucial for the early detection of cIVA, but it may not reliably protect against neonatal metabolic decompensations. This assumption is clearly confirmed by the results of the meta-analysis demonstrating showing a high frequency of neonatal decompensations in cIVA despite NBS (this meta-analysis 70% versus 54% in a recent observational study (4)). In a relevant number of individuals with cIVA (40-46% (4, 9)), acute metabolic decompensations occurred within hours to a few days after birth and, therefore, already before NBS results were available (8). Despite this, NBS might be partially benefical even for symptomatic individuals with cIVA since it often helps to shorten the path to diagnosis and specific treatment and thus helps to mitigate the negative consequences of neonatal metabolic decompensations (4).

However, as occurrence of EO decompensation was shown to have major impact on cognitive outcome in cIVA (4) and with timely NBS reports some EO metabolic decompensations can be prevented (8, 10), all efforts should be taken to optimized NBS processes for an as early as possible NBS report.

### Therapy and health care access

Another aspect that must be considered in this study is the question of the quality of treatment and care in different national helathcare systems, measured using the HAQ (41). In this meta-analysis, the HAQ showed no measurable effect on clinical and cognitive outcomes of individuals with cIVA identified by NBS versus diagnosed after onset of symptoms, but this should not mean that a high quality of treatment and care has no impact on the outcome. Rather, this could indicate that the current quality of treatment and the still high level of pre-diagnostic clinical manifestation (also in NBS) affect the outcome in general (42). This suggests that further improvements in care and treatment are needed to unleash the full potential of NBS.

### Limitations

A major limitation of meta-analyses is that they can only focus on the questions and hypotheses that have already been investigated in the included studies. Meta-analyses are designed to synthesize the results of existing research, but do not offer the opportunity to investigate new hypotheses. In addition, meta-analyses are highly dependent on the quality and design of the included studies, which can further limit the interpretation and generalizability of the results (43).

In this meta-analysis, several studies were excluded for specific reasons. Among the 20 publications included, not all could be utilized for every sub-analysis, as complete datasets were available from only a few studies. Additionally, many of the included studies had small sample sizes, with several being reports of case series that lacked systematic data reporting. Consequently, the risk of bias assessment for the included studies ranged from “some concerns” to “high concerns”.

It is also important to highlight that the follow-up periods for studies in the cIVA-Sympt group were longer than those in the cIVA-NBS group, particularly regarding mortality, neurological outcomes, and developmental delays. Furthermore, due to the small cohort size, individuals with the classic form of the disease, identified through HR screening were analyzed alongside the NBS cohort.

### Core outcome sets: Enhancing data standardization and greater comparability across studies

To reduce the exclusion of studies with lower data quality in future meta-analyses, we strongly encourage the use of international patient registries. These registries incorporate common data elements and standardized and semantically interoperable variables that clearly describe clinical and biochemical profiles, diagnostic approaches, treatment plans and follow-up procedures (44, 45). The establishment of standardized variables as core outcome sets is crucial to improve study comparability, especially in studies with individuals with rare diseases such as inborn metabolic disorders where patients numbers are limited. Such core outcome sets have already been developed for phenylketonuria and medium-chain acyl-CoA dehydrogenase deficiency in children through systematic reviews and Delphi surveys (46–48). To support this effort, we introduce a preliminary core outcome set for cIVA (Supplementary Table 3) based on the suggested core outcome set recently published for maple syrup urine disease (49).

## Conclusion

This meta-analysis shows that early diagnosis through NBS is essential for the timely initiation of therapy and for improving neurological outcomes and survival rates in individuals with cIVA even though it does not always prevent metabolic decompensations.

## Supporting information

Prisma Checklist

## Data Availability

Data availibilty statement
The analytical code can be made available upon request.

## Abbreviations

aIVA: attenuated IVA
C5: isovaleryl-carnitine
CI: confidence interval
cIVA: classic IVA
cIVA-NBS: classic IVA identified by NBS
cIVA-Sympt: classic IVA diagnosed after onset of symptoms
DDST: Denver developmental screening test
EO: early onset
HAQ: Health Quality Index
IVA: isovaleric aciduria
LO: late onset
NR: not reported
NBS: newborn screening
NH_3_: ammonia

## Funding

This study was generously supported by the Dietmar Hopp Foundation, St. Leon-Rot, Germany (grant number 1DH2011117 to G.F.H. and S.K.). The authors confirm independence from the sponsor; the content of the article has not been influenced by the sponsor.

## Ethics statement

For this article no studies with human or animal subjects were performed (evaluation of literature only). All procedures followed were in accordance with the ethical standards of the responsible committee on human experimentation (institutional and national) and with the Helsinki Declaration of 1975, as revised in 2000.

## Conflict of interest

S.K. and G.F.H. received research grants from the Dietmar Hopp Foundation, St. Leon-Rot, Germany (grant numbers 23011221, 1DH2011117). R.P. received consultancy fees from Immedica Pharma AB. The other authors have no conflicts of interest to disclose.

## Data availibilty statement

The analytical code can be made available upon request.

## Individual contributions

Conceptualization: A.T.R-H., S.K., U.M.; Data curation: A.T.R-H., F.G., E.S-B., R.P., M.Z., S.K., U.M.; Formal analysis: A.T.R-H., S.F.G.; Funding acquisition: S.K., G.F.H; Methodology: S.K., U.M., S.F.G., A.T.R-H.; Project administration: S.K., U.M.; Resources: S.K., G.F.H.; Software: S.F.G.; Supervision: S.K., U.M.; Validation: S.K., U.M., A.T.R-H.; Visualisation: S.F.G., A.T.R-H.; Writing-original draft: A.T.R-H., S.K., U.M.; Writing-review & editing: A.T.R-H., S.F.G., F.G., E.S-B., R.P., M.Z., G.F.H., S.K., U.M.

---

**Table:**
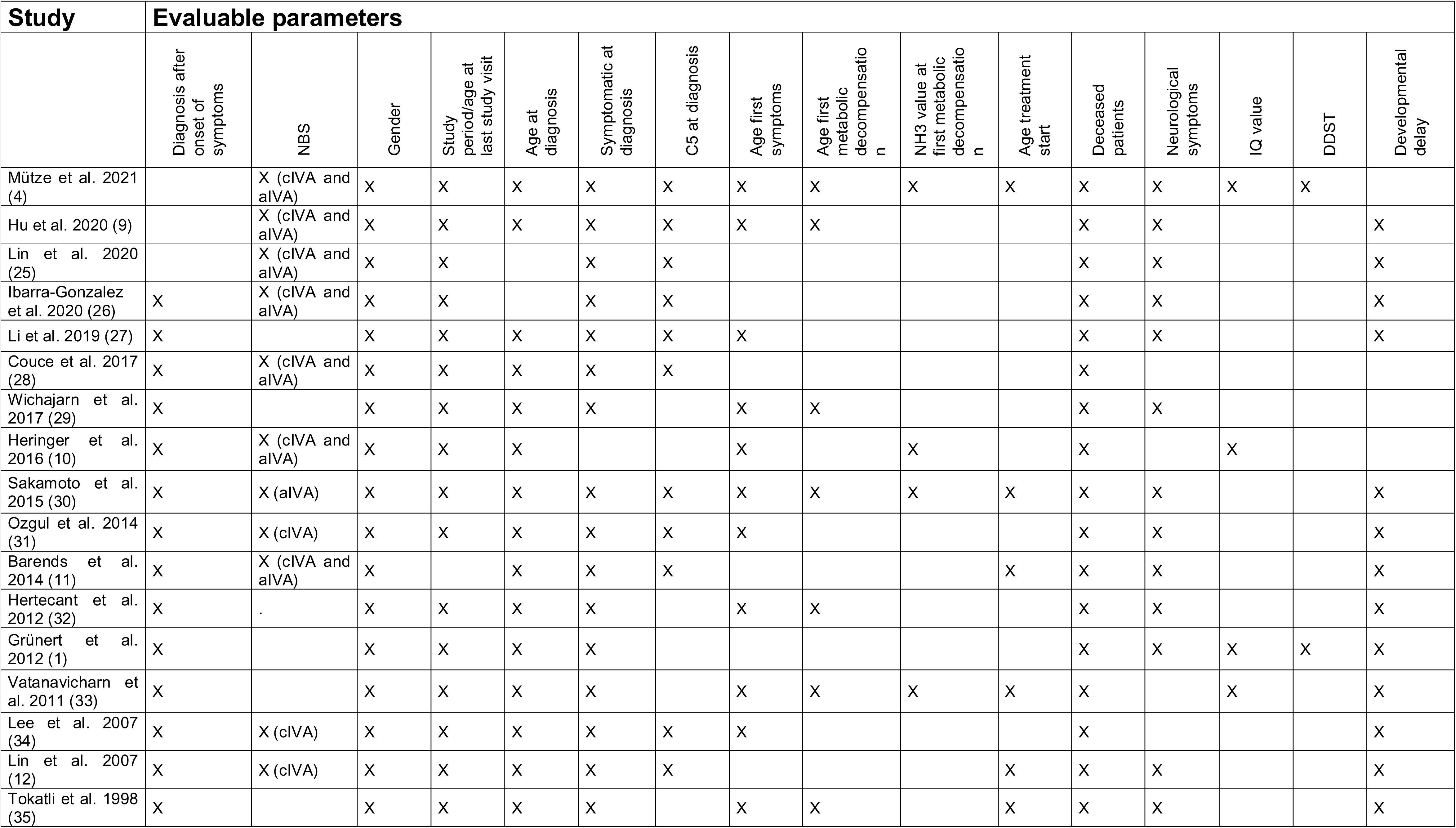
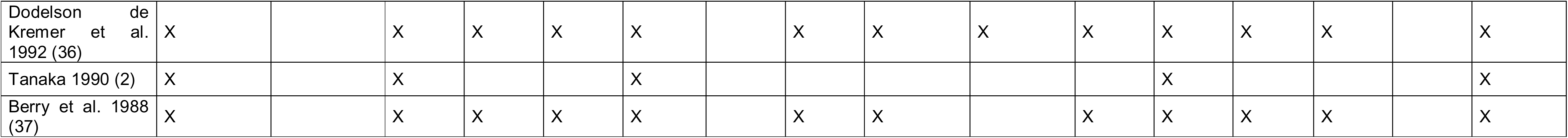
Evaluable parameters of studies included in the meta-analysis.

**Table: Evaluable parameters of studies included in the meta-analysis**
| Study | Evaluable parameters |  |  |  |  |  |  |  |  |  |  |  |  |  |  |  |
| --- | --- | --- | --- | --- | --- | --- | --- | --- | --- | --- | --- | --- | --- | --- | --- | --- |
|  | Diagnosis after onset of symptoms | NBS | Gender | Study period/age at last study visit | Age at diagnosis | Symptomatic at diagnosis | C5 at diagnosis | Age first symptoms | Age first metabolic decompensation | NH3 value at first metabolic decompensation | Age treatment start | Deceased patients | Neurological symptoms | IQ value | DDST | Developmental delay |
| Mütze et al. 2021 (4) |  | X (cIVA and aIVA) | X | X | X | X | X | X | X | X | X | X | X | X | X |  |
| Hu et al. 2020 (9) |  | X (cIVA and aIVA) | X | X | X | X | X | X | X |  |  | X | X |  |  | X |
| Lin et al. 2020 (25) |  | X (cIVA and aIVA) | X | X |  | X | X |  |  |  |  | X | X |  |  | X |
| Ibarra-Gonzalez et al. 2020 (26) | X | X (cIVA and aIVA) | X | X |  | X | X |  |  |  |  | X | X |  |  | X |
| Li et al. 2019 (27) | X |  | X | X | X | X | X | X |  |  |  | X | X |  |  | X |
| Couce et al. 2017 (28) | X | X (cIVA and aIVA) | X | X | X | X | X |  |  |  |  | X |  |  |  |  |
| Wichajarn et al. 2017 (29) | X |  | X | X | X | X |  | X | X |  |  | X | X |  |  |  |
| Heringer et al. 2016 (10) | X | X (cIVA and aIVA) | X | X | X |  |  | X |  | X |  | X |  | X |  |  |
| Sakamoto et al. 2015 (30) | X | X (aIVA) | X | X | X | X | X | X | X | X | X | X | X |  |  | X |
| Ozgul et al. 2014 (31) | X | X (cIVA) | X | X | X | X | X | X |  |  |  | X | X |  |  | X |
| Barends et al. 2014 (11) | X | X (cIVA and aIVA) | X |  | X | X | X |  |  |  | X | X | X |  |  | X |
| Hertecant et al. 2012 (32) | X | . | X | X | X | X |  | X | X |  |  | X | X |  |  | X |
| Grünert et al. 2012 (1) | X |  | X | X | X | X |  |  |  |  |  | X | X | X | X | X |
| Vatanavicharn et al. 2011 (33) | X |  | X | X | X | X |  | X | X | X | X | X |  | X |  | X |
| Lee et al. 2007 (34) | X | X (cIVA) | X | X | X | X | X | X |  |  |  | X |  |  |  | X |
| Lin et al. 2007 (12) | X | X (cIVA) | X | X | X | X | X |  |  |  | X | X | X |  |  | X |
| Tokatli et al. 1998 (35) | X |  | X | X | X | X |  | X | X |  | X | X | X |  |  | X |
| Dodelson de<br>Kremer et al.<br>1992 (36) | X |  | X | X | X | X |  | X | X | X | X | X | X | X |  | X |
| Tanaka 1990 (2) | X |  | X |  |  | X |  |  |  |  |  | X |  |  |  | X |
| Berry et al. 1988<br>(37) | X |  | X | X | X | X |  | X | X |  | X | X | X | X |  | X |

