## Supplementary Figure and Tables 1-3 for "Impact of newborn screening on survival and neurocognitive outcome in classic isovaleric aciduria: a meta-analysis"

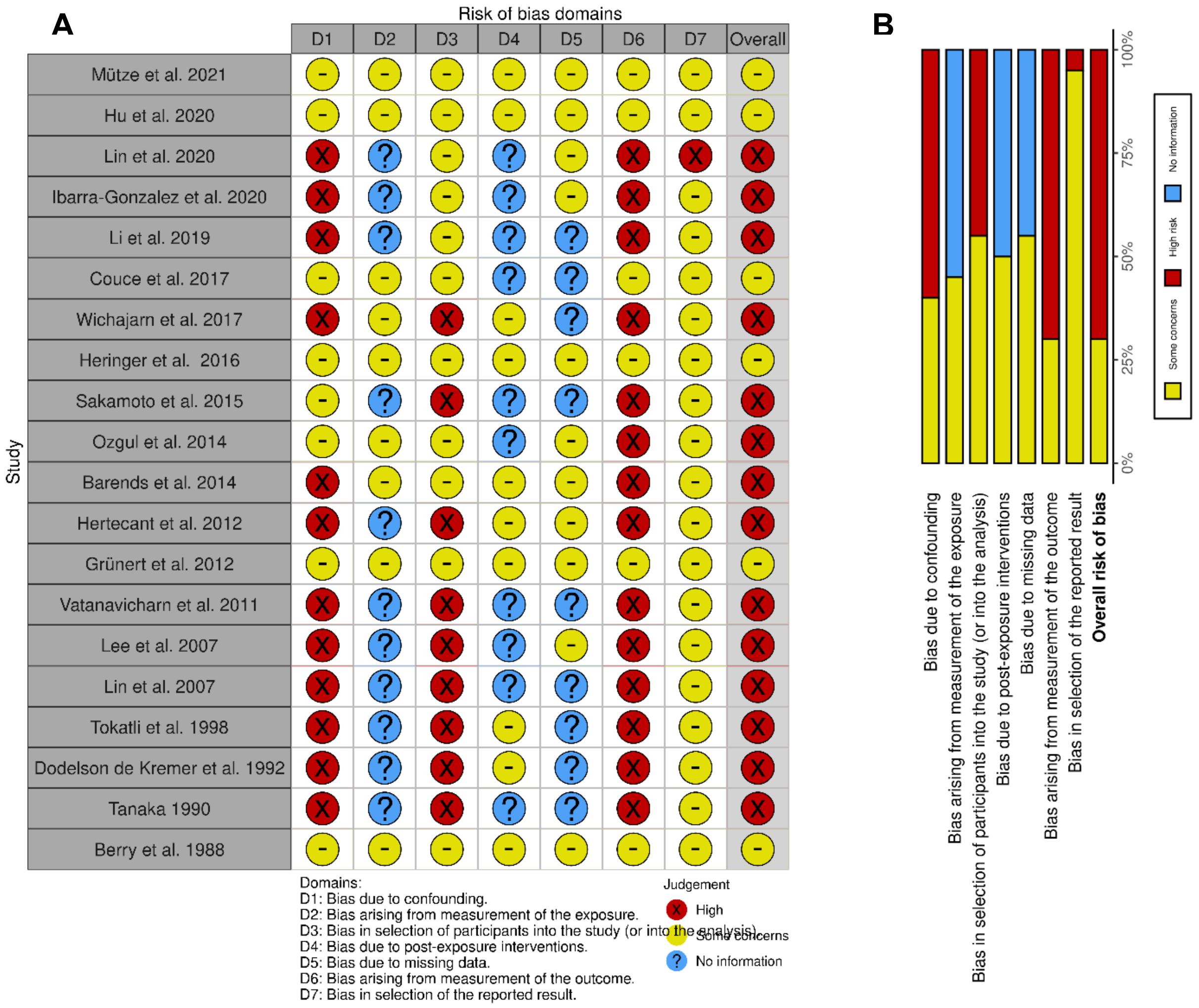


**Supplementary Figure: Risk-of-Bias assessment in non-randomized studies of exposures (ROBINS-E)** indicating the main seven domains (D1-D7) of risk-of-bias for each included study following as A) traffic light plot and B) summary plot (1). The Robvis risk-of bias tool (2) was used to create this figure.

**Supplementary Table 1: Included and excluded reports with reasons for exclusion (sorted in descending order by year of publication)**

| Authors | Publication Year | Journal | Title | Included in Meta-analysis (yes/no) | Comment |
|---|---|---|---|---|---|
| Bu et al. | 2022 | Zhonghua Er Ke Za Zhi | Disease spectrum analysis of children with inherited metabolic diseases detected by gas chromatography-mass spectrometry of urinary organic acids | no | - Report not retrieved (no access through the University of Heidelberg's library) |
| Mütze et al. | 2021 | J Inherit Metab Dis. | Newborn screening and disease variants predict neurological outcome in isovaleric aciduria | yes | - Study included in review<br>- Analysis of raw data set |
| Hu et al. | 2020 | Zhejiang Da Xue Xue Bao Yi Xue Ban | Screening and clinical analysis of isovaleric acidemia newborn in Zhejiang province | yes | - Study included in review<br>- Analysis of 14/15 patients because one patient could not be classified (authors contacted, no reply) |
| Szymańska et al. | 2020 | Diagnostics (Basel) | Long Term Follow-Up of Polish Patients with Isovaleric Aciduria. Clinical and Molecular Delineation of Isovaleric Aciduria | no | - Report excluded after record analysis<br>- Discrepancy between table and text (authors contacted, no reply) |
| Lin et al. | 2020 | Clin Chim Acta | Newborn screening for isovaleric acidemia in Quanzhou, China | yes | - Study included in review |
| Ibarra-González et al. | 2020 | Clin Chim Acta | Molecular analysis using targeted next generation DNA sequencing and clinical spectrum of Mexican patients with isovaleric acidemia | yes | - Study included in review<br>- Early onset defined until day 30, this classification used for patients in analysis |
| Li et al. | 2019 | Clin Chim Acta | Eight novel mutations detected from eight Chinese patients with isovaleric acidemia | yes | - Study included in review |
| Molema et al. | 2018 | J Inherit Metab Dis. | Fibroblast growth factor 21 as a biomarker for long-term complications in organic acidemias | no | - Report excluded after full text analysis<br>- No relevant outcome parameters |
| Keyfi et al. | 2018 | Hum Hered | Frequency of Inborn Errors of Metabolism in a Northeastern Iranian Sample with High Consanguinity Rates | no | - Report excluded after full text analysis<br>- No relevant outcome parameters |

| | | | | | |
|---|---|---|---|---|---|
| Schlune et al. | 2018 | Int J Neonatal Screen | Aspects of Newborn Screening in Isovaleric Acidemia | no | - Report excluded after full text analysis<br>- No relevant outcome parameters |
| Evans et al. | 2017 | J Pediatr | The Relationship between Dietary Intake, Growth, and Body Composition in Inborn Errors of Intermediary Protein Metabolism | no | - Report excluded after full text analysis<br>- No relevant outcome parameters |
| Couce et al. | 2017 | J Hum Genet | Genotype and phenotype characterization in a Spanish cohort with isovaleric acidemia | yes | - Study included in review<br>- Analysis of 15/16 patients because one patient was classified as family screening and asymptomatic at the age of 3 years |
| Wichajarn et al. | 2017 | Asian Biomedicine | Clinical and laboratory findings and outcomes of classic organic acidurias in children from north-eastern Thailand: a 5-year retrospective study | yes | - Study included in review |
| Heringer et al. | 2016 | J Inherit Metab Dis | Impact of age at onset and newborn screening on outcome in organic acidurias | yes | - Study included in review<br>- Analysis of raw data set<br>- Analysis of 50/83 patients because of exclusion of duplicates with analysis of Mütze et al. 2021 (N=23) and no possible classification (N=10) |
| Sakamoto et al. | 2015 | Tohoku J Exp Med | Phenotypic Variability and Newly Identified Mutations of the IVD Gene in Japanese Patients with Isovaleric Acidemia | yes | - Study included in review |
| Kölker et al. | 2015 | J Inherit Metab Dis. | The phenotypic spectrum of organic acidurias and urea cycle disorders. Part 2: the evolving clinical phenotype | no | - Report excluded after full text analysis<br>- Cohort overlap with Heringer et al. 2016 |
| Ozgul et al. | 2014 | Eur J Med Genet | Phenotypic and genotypic spectrum of Turkish patients with isovaleric acidemia | yes | - Study included in review |
| Barends et al. | 2014 | Molecular Genetics and Metabolism | Biochemical and molecular characteristics of patients with organic acidemias and urea | yes | - Study included in review |

| | | | | | |
|---|---|---|---|---|---|
| | | | cycle disorders identified through newborn screening | | |
| Nizon et al. | 2013 | Orphanet J Rare Dis | Long-term neurological outcome of a cohort of 80 patients with classical organic acidurias | no | - Report excluded after record analysis<br>- Focus on comparison of different disorders and only few data on IVA |
| Kaya et al. | 2013 | Gene | Identification of a novel IVD mutation in a consanguineous family with isovaleric acidemia | no | - Report excluded after record analysis<br>- Small cohort (N=3) and only few parameters |
| Hertecant et al. | 2012 | Eur J Med Genet | Clinical and molecular analysis of isovaleric acidemia patients in the United Arab Emirates reveals remarkable phenotypes and four novel mutations in the IVD gene | yes | - Study included in review<br>- Analysis of 6/7 patients because one patient could not be categorized |
| Dercksen et al. | 2012 | J Inherit Metab Dis. | Clinical variability of isovaleric acidemia in a genetically homogeneous population | no | - Report excluded after record analysis<br>- Difficulty in categorizing patients (authors contacted, no reply) |
| Grünert et al. | 2012 | Orphanet J Rare Dis | Clinical and neurocognitive outcome in symptomatic isovaleric acidemia | yes | - Study included in review<br>- "Late onset" in manuscript actually "late diagnosed"; 2 late-diagnosed: 1 unknown onset, 1 late onset<br>- Analysis of 19/21 patients because two patients could not be categorized |
| Vatanavicharn et al. | 2011 | Pediatr Int | Phenotypic and mutation spectrums of Thai patients with isovaleric acidemia | yes | - Study included in review |
| Kucuk et al. | 2010 | J Inherit Metab Dis. | Six novel mutations in Turkish patients with isovaleric acidemia | no | - Report not retrieved (no access through the University of Heidelberg's library) |
| Wasant et al. | 2008 | Clin Chim Acta | Organic acid disorders detected by urine organic acid analysis: twelve cases in Thailand over three-year experience | no | - Report excluded after record analysis<br>- Small cohort (N=3) and only few parameters |

| | | | | | |
|---|---|---|---|---|---|
| Lee et al. | 2007 | Mol Genet Metab | Different spectrum of mutations of isovaleryl-CoA dehydrogenase (IVD) gene in Korean patients with isovaleric acidemia | yes | - Study included in analysis<br>- Analysis of 5/7 patients because two patients could not be categorized |
| Joshi et al. | 2007 | Brain Dev | Clinical characteristics of neonates with inborn errors of metabolism detected by Tandem MS analysis in Oman | no | - Report excluded after record analysis<br>- Small cohort (N=3) and only few parameters |
| Loots et al. | 2007 | Eur J Clin Nutr | Amino-acid depletion induced by abnormal amino-acid conjugation and protein restriction in isovaleric acidemia | no | - Report excluded after full text analysis<br>- No relevant outcome parameters |
| Lin et al. | 2007 | Mol Genet Metab | Genetic mutation profile of isovaleric acidemia patients in Taiwan | yes | - Study included in analysis |
| Wasant et al. | 2006 | J Inherit Metab Dis. | Isovaleric acidemia in Thai infants: A report of 5 cases | no | - Report excluded after full text analysis<br>- Cohort overlap with Heringer et al. 2016 |
| Ensenauer et al. | 2004 | Am J Hum Genet | A common mutation is associated with a mild, potentially asymptomatic phenotype in patients with isovaleric acidemia diagnosed by newborn screening | no | - Report excluded after full text analysis<br>- No relevant outcome parameters |
| Wasant et al. | 1999 | Southeast Asian J Trop Med Public Health | Detection of inherited metabolic disorders via tandem mass spectrometry in Thai infants | no | - Report excluded after full text analysis<br>- No relevant outcome parameters |
| Wasant et al. | 1999 | Southeast Asian J Trop Med Public Health | Inherited metabolic disorders in Thailand-Siriraj experience | no | - Article not retrieved (no access through the University of Heidelberg's library) |
| Tokatli et al. | 1998 | Turk J Pediatr | Isovaleric acidemia. Clinical presentation of 6 cases | yes | - Study included in analysis |
| Yoshida et al. | 1998 | Ryoikibetsu Shokogun Shirizu | Isovaleric acidemia | no | - Article not retrieved (no access through the University of Heidelberg's library) |
| Itoh et al. | 1996 | Tohoku J Exp Med | Effect of carnitine administration on glycine metabolism in patients with isovaleric acidemia: significance of acetylcarnitine | no | - Report excluded after full text analysis<br>- No relevant outcome parameters |

| | | | | | |
|---|---|---|---|---|---|
| | | | determination to estimate the proper carnitine dose | | |
| Worthen et al. | 1994 | Brain Dev | Comparative frequency and severity of hypoglycemia in selected organic acidemias, branched chain amino acidemia, and disorders of fructose metabolism | no | - Report excluded after full text analysis<br>- No relevant outcome parameters |
| Kahler et al. | 1994 | J Pediatr | Pancreatitis in patients with organic acidemias | no | - Report excluded after full text analysis<br>- No relevant outcome parameters |
| Stanley et al. | 1993 | Pediatr Res | Renal handling of carnitine in secondary carnitine deficiency disorders | no | - Report excluded after full text analysis<br>- No relevant outcome parameters |
| Dodelson de Kremer et al. | 1992 | Medicina (B Aires) | Phenotypic expression variation of isovaleric acidemia in Argentinian patients. A long-term follow-up | yes | - Study included in analysis |
| Tanaka | 1990 | Prog Clin Biol Res | Isovaleric acidemia: personal history, clinical survey and study of the molecular basis | yes | - Study included in analysis |
| Gerdes et al. | 1989 | Ugeskr Laeger | Isovaleric acidemia | no | - Report excluded after record analysis<br>- Small cohort (N=3) and only few parameters |
| Berry et al. | 1988 | J Pediatr | Isovaleric acidemia: medical and neurodevelopmental effects of long-term therapy | yes | - Study included in analysis |
| Bakkeren et al. | 1982 | Tijdschr Kindergeneeskd | Isovaleric acidemia: identical biochemical picture in 3 patients with variable clinical manifestations | no | - Report excluded after record analysis<br>- Small cohort (N=3) and only few parameters |
| Prieto et al. | 1980 | Revista de investigacion clinica-clinical and translational investigation | The use of glycine in the acute management of isovaleric acidemia | no | - Article not retrieved (no access through the University of Heidelberg's library) |

| | | | | | |
|---|---|---|---|---|---|
| Saudubray et al. | 1976 | Arch Fr Pediatr | Isovaleric acidemia. Study and treatment in 3 brothers | no | - Report excluded after record analysis<br>- Small cohort (N=3) and only few parameters |
| Efron et al. | 1967 | Am J Dis Child | Isovaleric acidemia | no | - Report excluded after full text analysis<br>- No relevant outcome parameters |

### Supplementary Table 2: Results from the data collection

| Publication | Country | N Individuals in paper | N Individuals included in meta-analysis | HAQ | Diagnostic Mode | | General | | | | | | Diagnosis | | | | | | | Metabolic Decompensation | | | | | | Treatment | | | Mortality | | | | Outcome | | | | | | | | | | | | |
|---|---|---|---|---|---|---|---|---|---|---|---|---|---|---|---|---|---|---|---|---|---|---|---|---|---|---|---|---|---|---|---|---|---|---|---|---|---|---|---|---|---|---|---|---|---|
| | | | | Mean | | | N | N Male | N Female | Age last follow-up [months] N | Age last follow-up [months] Mean | Age last follow-up [months] SD | Age diagnosis [days] N | Age diagnosis [days] Mean | Age diagnosis [days] SD | N Symptomatic Diagnosis | C5 at diagnosis [µmol/L] N | C5 at diagnosis [µmol/L] Mean | C5 at diagnosis [µmol/L] SD | Age first metabolic decompensation [days] N | Age first metabolic decompensation [days] Mean | Age first metabolic decompensation [days] SD | NH3 first decompensation [µmol/L] N | NH3 first decompensation [µmol/L] Mean | NH3 first decompensation [µmol/L] SD | Age treatment start [days] N | Age treatment start [days] Mean | Age treatment start [days] SD | N Died | Age Death [months] N | Age Death [months] Mean | Age Death [months] SD | N Neurologic symptoms | N Developmental Delay | N DDST | N age-appropriate DDST | Age DDST [months] N | Age DDST [months] Mean | Age DDST [months] SD | Age IQ test [years] N | Age IQ test [years] Mean | Age IQ test [years] SD | IQ test [value] N | IQ test [value] Mean | IQ test [value] SD |
| Mütze et al. 2021 | Germany | 91 | 91 | 82,4 | NBS | total | 24 | 10 | 14 | 24 | 128,9 | 77,1 | 24 | 5,5 | 3,3 | NR | 23 | 11,3 | 4,6 | 17 | 174,8 | 331,0 | 17,0 | 376,1 | 413,7 | 22 | 5,4 | 3,5 | 0 | NR | NR | NR | 7 | NR | 11 | 9 | 11 | 51,4 | 39,9 | 16 | 10,3 | 6,0 | 16 | 90,7 | 10,1 |
| | | | | | NBS | EO | 13 | 3 | 10 | 13 | 87,0 | 60,0 | 13 | 5,1 | 2,3 | NR | 13 | 9,9 | 3,9 | 13 | 3,8 | 2,2 | 13,0 | 453,3 | 447,4 | 13 | 4,4 | 2,3 | 0 | NR | NR | NR | NR | NR | 9 | 7 | 9 | 44,5 | 23,7 | 8 | 7,3 | 4,6 | 8 | 85,5 | 9,9 |
| | | | | | NBS | LO | 6 | 4 | 2 | 6 | 177,5 | 37,9 | 6 | 3,2 | 3,8 | NR | 5 | 13,2 | 5,0 | 4 | 730,5 | 214,0 | 4,0 | 125,0 | 48,0 | 4 | 4,0 | 5,2 | 0 | NR | NR | NR | NR | NR | 0 | NR | NR | NR | NR | 5 | 13,2 | 4,8 | 5 | 93,8 | 6,8 |
| | | | | | NBS | asymptomatic | 5 | 3 | 2 | 5 | 179,9 | 95,3 | 5 | 9,4 | 1,5 | NR | 5 | 13,2 | 5,6 | NR | NR | NR | NR | NR | NR | 5 | 9,2 | 2,0 | 0 | NR | NR | NR | 0 | NR | 2 | 2 | 2 | 82,3 | 95,2 | 3 | 13,7 | 8,4 | 3 | 99,3 | 9,0 |
| | | | | | Sym pt. | total | 0 | NR | NR | NR | NR | NR | NR | NR | NR | NR | NR | NR | NR | NR | NR | NR | NR | NR | NR | NR | NR | NR | NR | NR | NR | NR | NR | NR | NR | NR | NR | NR | NR | NR | NR | NR | NR | NR | NR |
| | | | | | Sym pt. | EO | 0 | NR | NR | NR | NR | NR | NR | NR | NR | NR | NR | NR | NR | NR | NR | NR | NR | NR | NR | NR | NR | NR | NR | NR | NR | NR | NR | NR | NR | NR | NR | NR | NR | NR | NR | NR | NR | NR | NR |
| | | | | | Sym pt. | LO | 0 | NR | NR | NR | NR | NR | NR | NR | NR | NR | NR | NR | NR | NR | NR | NR | NR | NR | NR | NR | NR | NR | NR | NR | NR | NR | NR | NR | NR | NR | NR | NR | NR | NR | NR | NR | NR | NR | NR |
| Hu et al. 2020 | Zheijang, China | 15 | 14 | 72,1 | NBS | total | 10 | 5 | 5 | 9 | 18,2 | 17,9 | 10 | 19,8 | 6,3 | 4 | 10 | 9,9 | 2,4 | 3 | 7,7 | 5,0 | NR | NR | NR | NR | NR | NR | 1 | 1 | 0,2 | NA | 2 | 2 | NR | NR | NR | NR | NR | NR | NR | NR | NR | NR | NR |
| | | | | | NBS | EO | 3 | 2 | 1 | 3 | 20,1 | 23,5 | 3 | 18,7 | 8,1 | 3 | 3 | 10,7 | 0,8 | 3 | 7,7 | 5,0 | NR | NR | NR | NR | NR | NR | 1 | 1 | 0,2 | NA | 0 | 0 | NR | NR | NR | NR | NR | NR | NR | NR | NR | NR | NR |
| | | | | | NBS | LO | 2 | 2 | 0 | 2 | 35,5 | 14,8 | 2 | 19,0 | 2,8 | 1 | 2 | 6,8 | 0,9 | NR | NR | NR | NR | NR | NR | NR | NR | NR | 0 | NA | NA | NA | 2 | 2 | NR | NR | NR | NR | NR | NR | NR | NR | NR | NR | NR |
| | | | | | NBS | asymptomatic | 5 | 1 | 4 | 4 | 8,1 | 8,4 | 5 | 20,8 | 7,3 | 0 | 5 | 10,6 | 2,5 | NR | NR | NR | NR | NR | NR | NR | NR | NR | 0 | NA | NA | NA | 0 | 0 | NR | NR | NR | NR | NR | NR | NR | NR | NR | NR | NR |
| | | | | | Sym pt. | total | 0 | NR | NR | NR | NR | NR | NR | NR | NR | NR | NR | NR | NR | NR | NR | NR | NR | NR | NR | NR | NR | NR | NR | NR | NR | NR | NR | NR | NR | NR | NR | NR | NR | NR | NR | NR | NR | NR | NR |
| | | | | | Sym pt. | EO | 0 | NR | NR | NR | NR | NR | NR | NR | NR | NR | NR | NR | NR | NR | NR | NR | NR | NR | NR | NR | NR | NR | NR | NR | NR | NR | NR | NR | NR | NR | NR | NR | NR | NR | NR | NR | NR | NR | NR |
| | | | | | Sym pt. | LO | 0 | NR | NR | NR | NR | NR | NR | NR | NR | NR | NR | NR | NR | NR | NR | NR | NR | NR | NR | NR | NR | NR | NR | NR | NR | NR | NR | NR | NR | NR | NR | NR | NR | NR | NR | NR | NR | NR | NR |
| Lin et al. 2020 | | 5 | 5 | 72,1 | NBS | total | 1 | 1 | 0 | 1 | 0,4 | NR | NR | NR | NR | 1 | 1 | 7,2 | NR | NR | NR | NR | NR | NR | NR | NR | NR | NR | 1 | 1 | 0,4 | NR | 0 | 0 | NR | NR | NR | NR | NR | NR | NR | NR | NR | NR | NR |

| | | | | | | | | | | | | | | | | | | | | | | | | | | | | | | | | | | | | | | | | | | | | | |
|---|---|---|---|---|---|---|---|---|---|---|---|---|---|---|---|---|---|---|---|---|---|---|---|---|---|---|---|---|---|---|---|---|---|---|---|---|---|---|---|---|---|---|---|---|---|
| | Quanzhou, China | | | | NBS | EO | 1 | 1 | 0 | 1 | 0,4 | NR | NR | NR | NR | 1 | 1 | 7,2 | NR | NR | NR | NR | NR | NR | NR | NR | NR | NR | 1 | 1 | 0,4 | NR | 0 | 0 | NR | NR | NR | NR | NR | NR | NR | NR | NR | NR | NR |
| | | | | | NBS | LO | 0 | NR | NR | NR | NR | NR | NR | NR | NR | NR | NR | NR | NR | NR | NR | NR | NR | NR | NR | NR | NR | NR | NR | NR | NR | NR | NR | NR | NR | NR | NR | NR | NR | NR | NR | NR | NR | NR | NR |
| | | | | | NBS | asymptomatic | 0 | NR | NR | NR | NR | NR | NR | NR | NR | NR | NR | NR | NR | NR | NR | NR | NR | NR | NR | NR | NR | NR | NR | NR | NR | NR | NR | NR | NR | NR | NR | NR | NR | NR | NR | NR | NR | NR | NR |
| | | | | | Sym pt. | total | 0 | NR | NR | NR | NR | NR | NR | NR | NR | NR | NR | NR | NR | NR | NR | NR | NR | NR | NR | NR | NR | NR | NR | NR | NR | NR | NR | NR | NR | NR | NR | NR | NR | NR | NR | NR | NR | NR | NR |
| | | | | | Sym pt. | EO | 0 | NR | NR | NR | NR | NR | NR | NR | NR | NR | NR | NR | NR | NR | NR | NR | NR | NR | NR | NR | NR | NR | NR | NR | NR | NR | NR | NR | NR | NR | NR | NR | NR | NR | NR | NR | NR | NR | NR |
| | | | | | Sym pt. | LO | 0 | NR | NR | NR | NR | NR | NR | NR | NR | NR | NR | NR | NR | NR | NR | NR | NR | NR | NR | NR | NR | NR | NR | NR | NR | NR | NR | NR | NR | NR | NR | NR | NR | NR | NR | NR | NR | NR | NR |
| Ibarra-Gonzalez et al. 2020 | Mexico | 13 | 13 | 59,9 | NBS | total | 0 | NR | NR | NR | NR | NR | NR | NR | NR | NR | NR | NR | NR | NR | NR | NR | NR | NR | NR | NR | NR | NR | NR | NR | NR | NR | NR | NR | NR | NR | NR | NR | NR | NR | NR | NR | NR | NR | NR |
| | | | | | NBS | EO | 0 | NR | NR | NR | NR | NR | NR | NR | NR | NR | NR | NR | NR | NR | NR | NR | NR | NR | NR | NR | NR | NR | NR | NR | NR | NR | NR | NR | NR | NR | NR | NR | NR | NR | NR | NR | NR | NR | NR |
| | | | | | NBS | LO | 0 | NR | NR | NR | NR | NR | NR | NR | NR | NR | NR | NR | NR | NR | NR | NR | NR | NR | NR | NR | NR | NR | NR | NR | NR | NR | NR | NR | NR | NR | NR | NR | NR | NR | NR | NR | NR | NR | NR |
| | | | | | NBS | asymptomatic | 0 | NR | NR | NR | NR | NR | NR | NR | NR | NR | NR | NR | NR | NR | NR | NR | NR | NR | NR | NR | NR | NR | NR | NR | NR | NR | NR | NR | NR | NR | NR | NR | NR | NR | NR | NR | NR | NR | NR |
| | | | | | Sym pt. | total | 10 | 5 | 5 | 10 | 181,2 | 50,1 | NR | NR | NR | 10 | 10 | 8,8 | 4,5 | NR | NR | NR | NR | NR | NR | NR | NR | NR | 0 | NR | NR | NR | 8 | 8 | NR | NR | NR | NR | NR | NR | NR | NR | NR | NR | NR |
| | | | | | Sym pt. | EO | 6 | 3 | 3 | 6 | 188,0 | 61,5 | NR | NR | NR | 6 | 6 | 8,7 | 2,7 | NR | NR | NR | NR | NR | NR | NR | NR | NR | 0 | NR | NR | NR | 6 | 6 | NR | NR | NR | NR | NR | NR | NR | NR | NR | NR | NR |
| | | | | | Sym pt. | LO | 4 | 2 | 2 | 4 | 171,0 | 31,6 | NR | NR | NR | 4 | 4 | 8,9 | 7,0 | NR | NR | NR | NR | NR | NR | NR | NR | NR | 0 | NR | NR | NR | 2 | 2 | NR | NR | NR | NR | NR | NR | NR | NR | NR | NR | NR |
| Li et al. 2019 | Peking, China | 8 | 8 | 69,2 | NBS | total | 0 | NR | NR | NR | NR | NR | NR | NR | NR | NR | NR | NR | NR | NR | NR | NR | NR | NR | NR | NR | NR | NR | NR | NR | NR | NR | NR | NR | NR | NR | NR | NR | NR | NR | NR | NR | NR | NR | NR |
| | | | | | NBS | EO | 0 | NR | NR | NR | NR | NR | NR | NR | NR | NR | NR | NR | NR | NR | NR | NR | NR | NR | NR | NR | NR | NR | NR | NR | NR | NR | NR | NR | NR | NR | NR | NR | NR | NR | NR | NR | NR | NR | NR |
| | | | | | NBS | LO | 0 | NR | NR | NR | NR | NR | NR | NR | NR | NR | NR | NR | NR | NR | NR | NR | NR | NR | NR | NR | NR | NR | NR | NR | NR | NR | NR | NR | NR | NR | NR | NR | NR | NR | NR | NR | NR | NR | NR |
| | | | | | NBS | asymptomatic | 0 | NR | NR | NR | NR | NR | NR | NR | NR | NR | NR | NR | NR | NR | NR | NR | NR | NR | NR | NR | NR | NR | NR | NR | NR | NR | NR | NR | NR | NR | NR | NR | NR | NR | NR | NR | NR | NR | NR |
| | | | | | Sym pt. | total | 8 | 3 | 5 | 8 | 70,5 | 77,0 | 6 | 894,5 | 831,3 | 8 | 8 | 9,7 | 4,8 | NR | NR | NR | NR | NR | NR | NR | NR | NR | 2 | 2 | 0,4 | 0,1 | 2 | 2 | NR | NR | NR | NR | NR | NR | NR | NR | NR | NR | NR |
| | | | | | Sym pt. | EO | 5 | 2 | 3 | 5 | 42,2 | 45,5 | 3 | 388,3 | 316,1 | 5 | 5 | 11,5 | 5,2 | NR | NR | NR | NR | NR | NR | NR | NR | NR | 2 | 2 | 0,4 | 0,1 | 2 | 2 | NR | NR | NR | NR | NR | NR | NR | NR | NR | NR | NR |
| | | | | | Sym pt. | LO | 3 | 1 | 2 | 3 | 117,7 | 106,2 | 3 | 1400,7 | 929,4 | 3 | 3 | 6,6 | 1,8 | NR | NR | NR | NR | NR | NR | NR | NR | NR | 0 | NR | NR | NR | 0 | 0 | NR | NR | NR | NR | NR | NR | NR | NR | NR | NR | NR |
| Couce et al. 2017 | Spain | 16 | 15 | 82,3 | NBS | total | 2 | 0 | 2 | 2 | 70,0 | 90,5 | 2 | 4,0 | 2,8 | 1 | 2 | 11,8 | 5,5 | NR | NR | NR | NR | NR | NR | NR | NR | NR | 0 | NR | NR | NR | NR | NR | NR | NR | NR | NR | NR | NR | NR | NR | 2 | 85,0 | 0,0 |
| | | | | | NBS | EO | 1 | 0 | 1 | 1 | 6,0 | NA | 1 | 6,0 | NR | 1 | 1 | 7,9 | NR | NR | NR | NR | NR | NR | NR | NR | NR | NR | 0 | NR | NR | NR | NR | NR | NR | NR | NR | NR | NR | NR | NR | NR | 1 | 85,0 | NA |
| | | | | | NBS | LO | 0 | NR | NR | NR | NR | NR | NR | NR | NR | NR | NR | NR | NR | NR | NR | NR | NR | NR | NR | NR | NR | NR | NR | NR | NR | NR | NR | NR | NR | NR | NR | NR | NR | NR | NR | NR | NR | NR | NR |
| | | | | | NBS | asymptomatic | 1 | 0 | 1 | 1 | 134,0 | NA | 1 | 2,0 | NR | 0 | 1 | 15,7 | NR | NR | NR | NR | NR | NR | NR | NR | NR | NR | 0 | NR | NR | NR | NR | NR | NR | NR | NR | NR | NR | NR | NR | NR | 1 | 85,0 | NR |
| | | | | | Sym pt. | total | 6 | 3 | 3 | 6 | 152,7 | 106,8 | 6 | 613,8 | 847,3 | 6 | 4 | 8,1 | 5,8 | NR | NR | NR | NR | NR | NR | NR | NR | NR | 1 | 1 | 0,3 | NR | NR | NR | NR | NR | NR | NR | NR | NR | NR | NR | 5 | 87,4 | 2,9 |
| | | | | | Sym pt. | EO | 3 | 2 | 1 | 3 | 67,4 | 58,6 | 3 | 17,7 | 10,7 | 3 | 2 | 10,1 | 8,8 | NR | NR | NR | NR | NR | NR | NR | NR | NR | 1 | 1 | 0,3 | NR | NR | NR | NR | NR | NR | NR | NR | NR | NR | NR | 2 | 86,0 | 1,4 |
| | | | | | Sym pt. | LO | 3 | 1 | 2 | 3 | 238,0 | 56,5 | 3 | 1210,0 | 835,5 | 3 | 2 | 6,1 | 2,5 | NR | NR | NR | NR | NR | NR | NR | NR | NR | 0 | NR | NR | NR | NR | NR | NR | NR | NR | NR | NR | NR | NR | NR | 3 | 88,3 | 3,5 |
| Wichajarn et al. 2017 | Thailand | 5 | 5 | 69,6 | NBS | total | 0 | NR | NR | NR | NR | NR | NR | NR | NR | NR | NR | NR | NR | NR | NR | NR | NR | NR | NR | NR | NR | NR | NR | NR | NR | NR | NR | NR | NR | NR | NR | NR | NR | NR | NR | NR | NR | NR | NR |
| | | | | | NBS | EO | 0 | NR | NR | NR | NR | NR | NR | NR | NR | NR | NR | NR | NR | NR | NR | NR | NR | NR | NR | NR | NR | NR | NR | NR | NR | NR | NR | NR | NR | NR | NR | NR | NR | NR | NR | NR | NR | NR | NR |

| | | | | | | | | | | | | | | | | | | | | | | | | | | | | | | | | | | | | | | | | | | | | | |
|---|---|---|---|---|---|---|---|---|---|---|---|---|---|---|---|---|---|---|---|---|---|---|---|---|---|---|---|---|---|---|---|---|---|---|---|---|---|---|---|---|---|---|---|---|---|
| | | | | | NBS | LO | 0 | NR | NR | NR | NR | NR | NR | NR | NR | NR | NR | NR | NR | NR | NR | NR | NR | NR | NR | NR | NR | NR | NR | NR | NR | NR | NR | NR | NR | NR | NR | NR | NR | NR | NR | NR | NR | NR | NR |
| | | | | | NBS | asymptomatic | 0 | NR | NR | NR | NR | NR | NR | NR | NR | NR | NR | NR | NR | NR | NR | NR | NR | NR | NR | NR | NR | NR | NR | NR | NR | NR | NR | NR | NR | NR | NR | NR | NR | NR | NR | NR | NR | NR | NR |
| | | | | | Sym pt. | total | 5 | 2 | 3 | 5 | 21,9 | 19,3 | 5 | 18,2 | 6,0 | 5 | NR | NR | NR | 5 | 6,6 | 4,4 | NR | NR | NR | NR | NR | NR | 1 | 1 | 0,6 | NR | 0 | NR | NR | NR | NR | NR | NR | NR | NR | NR | NR | NR | NR |
| | | | | | Sym pt. | EO | 5 | 2 | 3 | 5 | 21,9 | 19,3 | 5 | 18,2 | 6,0 | 5 | NR | NR | NR | 5 | 6,6 | 4,4 | NR | NR | NR | NR | NR | NR | 1 | 1 | 0,6 | NR | 0 | NR | NR | NR | NR | NR | NR | NR | NR | NR | NR | NR | NR |
| | | | | | Sym pt. | LO | 0 | NR | NR | NR | NR | NR | NR | NR | NR | NR | NR | NR | NR | NR | NR | NR | NR | NR | NR | NR | NR | NR | NR | NR | NR | NR | NR | NR | NR | NR | NR | NR | NR | NR | NR | NR | NR | NR | NR |
| Heringer et al. 2016 | EIMD | 83 | 50 | 84,6 | NBS | total | 13 | 2 | 11 | 13 | 120,4 | 81,4 | 13 | 6,8 | 7,5 | NR | NR | NR | NR | NR | NR | NR | 4 | 343,0 | 445,5 | NR | NR | NR | 0 | NR | NR | NR | NR | NR | NR | NR | NR | NR | NR | NR | NR | NR | NR | NR | NR |
| | | | | | NBS | EO | 7 | 1 | 6 | 7 | 125,9 | 84,6 | 7 | 11,3 | 7,7 | NR | NR | NR | NR | NR | NR | NR | 4 | 343,0 | 445,5 | NR | NR | NR | 0 | NR | NR | NR | NR | NR | NR | NR | NR | NR | NR | NR | NR | NR | NR | NR | NR |
| | | | | | NBS | LO | 3 | 0 | 3 | 3 | 137,1 | 107,3 | 3 | 1,0 | 0,0 | NR | NR | NR | NR | NR | NR | NR | NR | NR | NR | NR | NR | NR | 0 | NR | NR | NR | NR | NR | NR | NR | NR | NR | NR | NR | NR | NR | NR | NR | NR |
| | | | | | NBS | asymptomatic | 3 | 1 | 2 | 3 | 91,0 | 70,3 | 3 | 2,3 | 2,5 | NR | NR | NR | NR | NR | NR | NR | NR | NR | NR | NR | NR | NR | 0 | NR | NR | NR | NR | NR | NR | NR | NR | NR | NR | NR | NR | NR | NR | NR | NR |
| | | | | | Sym pt. | total | 34 | 23 | 11 | 34 | 168,9 | 115,0 | 34 | 993,0 | 1489,3 | NR | NR | NR | NR | NR | NR | NR | 17 | 153,7 | 93,3 | NR | NR | NR | 0 | NR | NR | NR | NR | NR | NR | NR | NR | NR | NR | 10 | 16,0 | 12,1 | 10 | 86,5 | 25,9 |
| | | | | | Sym pt. | EO | 17 | 11 | 6 | 17 | 187,0 | 122,5 | 17 | 216,5 | 591,2 | NR | NR | NR | NR | NR | NR | NR | 9 | 191,0 | 84,9 | NR | NR | NR | 0 | NR | NR | NR | NR | NR | NR | NR | NR | NR | NR | 5 | 25,2 | 10,4 | 5 | 89,8 | 27,0 |
| | | | | | Sym pt. | LO | 17 | 12 | 5 | 17 | 150,8 | 107,5 | 17 | 1769,4 | 1715,9 | NR | NR | NR | NR | NR | NR | NR | 8 | 111,6 | 88,5 | NR | NR | NR | 0 | NR | NR | NR | NR | NR | NR | NR | NR | NR | NR | 5 | 6,8 | 3,2 | 5 | 83,2 | 27,4 |
| Sakamoto et al. 2015 | Japan | 5 | 5 | 88,4 | NBS | total | 0 | NR | NR | NR | NR | NR | NR | NR | NR | NR | NR | NR | NR | NR | NR | NR | NR | NR | NR | NR | NR | NR | NR | NR | NR | NR | NR | NR | NR | NR | NR | NR | NR | NR | NR | NR | NR | NR | NR |
| | | | | | NBS | EO | 0 | NR | NR | NR | NR | NR | NR | NR | NR | NR | NR | NR | NR | NR | NR | NR | NR | NR | NR | NR | NR | NR | NR | NR | NR | NR | NR | NR | NR | NR | NR | NR | NR | NR | NR | NR | NR | NR | NR |
| | | | | | NBS | LO | 0 | NR | NR | NR | NR | NR | NR | NR | NR | NR | NR | NR | NR | NR | NR | NR | NR | NR | NR | NR | NR | NR | NR | NR | NR | NR | NR | NR | NR | NR | NR | NR | NR | NR | NR | NR | NR | NR | NR |
| | | | | | NBS | asymptomatic | 0 | NR | NR | NR | NR | NR | NR | NR | NR | NR | NR | NR | NR | NR | NR | NR | NR | NR | NR | NR | NR | NR | NR | NR | NR | NR | NR | NR | NR | NR | NR | NR | NR | NR | NR | NR | NR | NR | NR |
| | | | | | Sym pt. | total | 4 | NR | NR | 3 | 44,5 | 37,7 | 4 | 85,5 | 92,7 | 4 | 3 | 7,4 | 2,2 | 4 | 893,0 | 1064,7 | 3,0 | 358,3 | 383,5 | 4 | 893,0 | 1064,7 | 0 | NR | NR | NR | 2 | 2 | NR | NR | NR | NR | NR | NR | NR | NR | NR | NR | NR |
| | | | | | Sym pt. | EO | 2 | NR | NR | 2 | 1,6 | NA | 2 | 6,0 | 5,7 | 2 | 2 | 8,7 | 0,5 | 2 | 6,0 | 5,7 | 2,0 | 482,5 | 449,0 | 2 | 6,0 | 5,7 | 0 | NR | NR | NR | 1 | 1 | NR | NR | NR | NR | NR | NR | NR | NR | NR | NR | NR |
| | | | | | Sym pt. | LO | 2 | NR | NR | 2 | 66,0 | 8,5 | 2 | 165,0 | 21,2 | 2 | 1 | 4,9 | NA | 2 | 1780,0 | 503,5 | 1,0 | 110,0 | NA | 2 | 1780,0 | 503,5 | 0 | NR | NR | NR | 1 | 1 | NR | NR | NR | NR | NR | NR | NR | NR | NR | NR | NR |
| Ozgul et al. 2014 | Turkey | 20 | 19 | 64,7 | NBS | total | 1 | 0 | 1 | 1 | 16,0 | NR | 1 | 7,0 | NR | 0 | 1 | 9,1 | NR | NR | NR | NR | NR | NR | NR | NR | NR | NR | NR | NR | NR | NR | NR | NR | NR | NR | NR | NR | NR | NR | NR | NR | NR | NR | NR |
| | | | | | NBS | EO | 0 | NR | NR | NR | NR | NR | NR | NR | NR | NR | NR | NR | NR | NR | NR | NR | NR | NR | NR | NR | NR | NR | NR | NR | NR | NR | NR | NR | NR | NR | NR | NR | NR | NR | NR | NR | NR | NR | NR |
| | | | | | NBS | LO | 0 | NR | NR | NR | NR | NR | NR | NR | NR | NR | NR | NR | NR | NR | NR | NR | NR | NR | NR | NR | NR | NR | NR | NR | NR | NR | NR | NR | NR | NR | NR | NR | NR | NR | NR | NR | NR | NR | NR |
| | | | | | NBS | asymptomatic | 1 | 0 | 1 | 1 | 16,0 | NR | 1 | 7,0 | NR | 0 | 1 | 9,1 | NR | NR | NR | NR | NR | NR | NR | NR | NR | NR | 0 | NR | NR | NR | 0 | 0 | NR | NR | NR | NR | NR | NR | NR | NR | NR | NR | NR |
| | | | | | Sym pt. | total | 18 | 8 | 10 | 16 | 129,4 | 85,0 | 18 | 882,2 | 1065,8 | 18 | 12 | 10,1 | 10,2 | NR | NR | NR | NR | NR | NR | NR | NR | NR | 2 | NR | NR | NR | 8 | 8 | NR | NR | NR | NR | NR | NR | NR | NR | NR | NR | NR |
| | | | | | Sym pt. | EO | 11 | 6 | 5 | 9 | 104,3 | 70,3 | 11 | 513,6 | 1100,6 | 11 | 8 | 12,3 | 11,8 | NR | NR | NR | NR | NR | NR | NR | NR | NR | 2 | NR | NR | NR | 5 | 5 | NR | NR | NR | NR | NR | NR | NR | NR | NR | NR | NR |
| | | | | | Sym pt. | LO | 7 | 2 | 5 | 7 | 161,6 | 96,4 | 7 | 1461,4 | 747,7 | 7 | 4 | 5,7 | 4,1 | NR | NR | NR | NR | NR | NR | NR | NR | NR | 0 | NR | NR | NR | 3 | 3 | NR | NR | NR | NR | NR | NR | NR | NR | NR | NR | NR |
| Barends et al. 2014 | Australia | 8 | 8 | 84,5 | NBS | total | 4 | 1 | 3 | NR | NR | NR | NR | NR | NR | 1 | 4 | 42,7 | 39,5 | NR | NR | NR | NR | NR | NR | 1 | NR | NR | 0 | NR | NR | NR | 0 | 0 | NR | NR | NR | NR | NR | NR | NR | NR | NR | NR | NR |
| | | | | | NBS | EO | 1 | 0 | 1 | NR | NR | NR | NR | NR | NR | 1 | 1 | 87,0 | NR | NR | NR | NR | NR | NR | NR | NR | NR | NR | 0 | NR | NR | NR | 0 | 0 | NR | NR | NR | NR | NR | NR | NR | NR | NR | NR | NR |
| | | | | | NBS | LO | 0 | NA | NA | NR | NR | NR | NR | NR | NR | NR | NR | NR | NR | NR | NR | NR | NR | NR | NR | NR | NR | NR | NR | NR | NR | NR | NR | NR | NR | NR | NR | NR | NR | NR | NR | NR | NR | NR | NR |

| | | | | | | | | | | | | | | | | | | | | | | | | | | | | | | | | | | | | | | | | | | | | | |
|---|---|---|---|---|---|---|---|---|---|---|---|---|---|---|---|---|---|---|---|---|---|---|---|---|---|---|---|---|---|---|---|---|---|---|---|---|---|---|---|---|---|---|---|---|---|
| | | | | | NBS | asymptomatic | 3 | 1 | 2 | NR | NR | NR | NR | NR | NR | 0 | 3 | 27,9 | 32,2 | NA | NR | NR | NR | NR | NR | 1 | 1,0 | NA | 0 | NR | NR | NR | 0 | 0 | NR | NR | NR | NR | NR | NR | NR | NR | NR | NR | NR |
| | | | | | Sym pt. | total | 2 | 1 | 1 | NR | NR | NR | 1 | 1068,0 | NR | 2 | 1 | 6,7 | NR | NR | NR | NR | NR | NR | NR | NR | NR | NR | 1 | NR | NR | NR | 1 | 1 | NR | NR | NR | NR | NR | NR | NR | NR | NR | NR | NR |
| | | | | | Sym pt. | EO | 1 | 0 | 1 | NR | NR | NR | NR | NR | NR | 1 | NR | NR | NR | NR | NR | NR | NR | NR | NR | NR | NR | NR | 1 | NR | NR | NR | 0 | 0 | NR | NR | NR | NR | NR | NR | NR | NR | NR | NR | NR |
| | | | | | Sym pt. | LO | 1 | 1 | 0 | NR | NR | NR | 1 | 1068,0 | NR | 1 | 1 | 6,7 | NR | NR | NR | NR | NR | NR | NR | NR | NR | NR | 0 | NR | NR | NR | 1 | 1 | NR | NR | NR | NR | NR | NR | NR | NR | NR | NR | NR |
| Hertecant et al. 2012 | United Arab Emirates | 7 | 6 | 66,5 | NBS | total | 0 | NR | NR | NR | NR | NR | NR | NR | NR | NR | NR | NR | NR | NR | NR | NR | NR | NR | NR | NR | NR | NR | NR | NR | NR | NR | NR | NR | NR | NR | NR | NR | NR | NR | NR | NR | NR | NR | NR | NR |
| | | | | | NBS | EO | 0 | NR | NR | NR | NR | NR | NR | NR | NR | NR | NR | NR | NR | NR | NR | NR | NR | NR | NR | NR | NR | NR | NR | NR | NR | NR | NR | NR | NR | NR | NR | NR | NR | NR | NR | NR | NR | NR | NR | NR |
| | | | | | NBS | LO | 0 | NR | NR | NR | NR | NR | NR | NR | NR | NR | NR | NR | NR | NR | NR | NR | NR | NR | NR | NR | NR | NR | NR | NR | NR | NR | NR | NR | NR | NR | NR | NR | NR | NR | NR | NR | NR | NR | NR | NR |
| | | | | | NBS | asymptomatic | 0 | NR | NR | NR | NR | NR | NR | NR | NR | NR | NR | NR | NR | NR | NR | NR | NR | NR | NR | NR | NR | NR | NR | NR | NR | NR | NR | NR | NR | NR | NR | NR | NR | NR | NR | NR | NR | NR | NR | NR |
| | | | | | Sym pt. | total | 6 | 3 | 3 | 6 | 106,0 | 57,5 | 4 | 457,3 | 881,8 | 6 | NR | NR | NR | 4 | 674,5 | 844,8 | NR | NR | NR | NR | NR | NR | 0 | NR | NR | NR | 2 | 2 | NR | NR | NR | NR | NR | NR | NR | NR | NR | NR | NR |
| | | | | | Sym pt. | EO | 4 | 2 | 2 | 4 | 118,0 | 68,1 | 3 | 16,3 | 5,0 | 4 | NR | NR | NR | 2 | 14,0 | 4,2 | NR | NR | NR | NR | NR | NR | 0 | NR | NR | NR | 2 | 2 | NR | NR | NR | NR | NR | NR | NR | NR | NR | NR | NR |
| | | | | | Sym pt. | LO | 2 | 1 | 1 | 2 | 84,0 | 33,9 | 1 | 1780,0 | NA | 2 | NR | NR | NR | 2 | 1335,0 | 629,3 | NR | NR | NR | NR | NR | NR | 0 | NR | NR | NR | 0 | 0 | NR | NR | NR | NR | NR | NR | NR | NR | NR | NR | NR |
| Grünert et al. 2012 | Germany | 21 | 19 | 76,9 | NBS | total | 0 | NR | NR | NR | NR | NR | NR | NR | NR | NR | NR | NR | NR | NR | NR | NR | NR | NR | NR | NR | NR | NR | NR | NR | NR | NR | NR | NR | NR | NR | NR | NR | NR | NR | NR | NR | NR | NR | NR | NR |
| | | | | | NBS | EO | 0 | NR | NR | NR | NR | NR | NR | NR | NR | NR | NR | NR | NR | NR | NR | NR | NR | NR | NR | NR | NR | NR | NR | NR | NR | NR | NR | NR | NR | NR | NR | NR | NR | NR | NR | NR | NR | NR | NR | NR |
| | | | | | NBS | LO | 0 | NR | NR | NR | NR | NR | NR | NR | NR | NR | NR | NR | NR | NR | NR | NR | NR | NR | NR | NR | NR | NR | NR | NR | NR | NR | NR | NR | NR | NR | NR | NR | NR | NR | NR | NR | NR | NR | NR | NR |
| | | | | | NBS | asymptomatic | 0 | NR | NR | NR | NR | NR | NR | NR | NR | NR | NR | NR | NR | NR | NR | NR | NR | NR | NR | NR | NR | NR | NR | NR | NR | NR | NR | NR | NR | NR | NR | NR | NR | NR | NR | NR | NR | NR | NR | NR |
| | | | | | Sym pt. | total | 19 | 10 | 9 | 19 | 139,4 | 90,0 | 19 | 559,4 | 834,2 | 19 | NR | NR | NR | NR | NR | NR | NR | NR | NR | NR | NR | NR | 1 | 1 | 0,5 | NA | 7 | 6 | 2 | 1 | 2 | 36,0 | 13,6 | 15 | 10,7 | 5,3 | 15 | 99,8 | 21,0 |
| | | | | | Sym pt. | EO | 19 | 10 | 9 | 19 | 139,4 | 90,0 | 19 | 559,4 | 834,2 | 19 | NR | NR | NR | NR | NR | NR | NR | NR | NR | NR | NR | NR | 1 | 1 | 0,5 | NA | 7 | 6 | 2 | 1 | 2 | 36,0 | 13,6 | 15 | 10,7 | 5,3 | 15 | 99,8 | 21,0 |
| | | | | | Sym pt. | LO | 0 | NR | NR | NR | NR | NR | NR | NR | NR | NR | NR | NR | NR | NR | NR | NR | NR | NR | NR | NR | NR | NR | NR | NR | NR | NR | NR | NR | NR | NR | NR | NR | NR | NR | NR | NR | NR | NR | NR | NR |
| Vatanavicharn et al. 2011 | Thailand | 5 | 5 | 60 | NBS | total | 0 | NR | NR | NR | NR | NR | NR | NR | NR | NR | NR | NR | NR | NR | NR | NR | NR | NR | NR | NR | NR | NR | NR | NR | NR | NR | NR | NR | NR | NR | NR | NR | NR | NR | NR | NR | NR | NR | NR | NR |
| | | | | | NBS | EO | 0 | NR | NR | NR | NR | NR | NR | NR | NR | NR | NR | NR | NR | NR | NR | NR | NR | NR | NR | NR | NR | NR | NR | NR | NR | NR | NR | NR | NR | NR | NR | NR | NR | NR | NR | NR | NR | NR | NR | NR |
| | | | | | NBS | LO | 0 | NR | NR | NR | NR | NR | NR | NR | NR | NR | NR | NR | NR | NR | NR | NR | NR | NR | NR | NR | NR | NR | NR | NR | NR | NR | NR | NR | NR | NR | NR | NR | NR | NR | NR | NR | NR | NR | NR | NR |
| | | | | | NBS | asymptomatic | 0 | NR | NR | NR | NR | NR | NR | NR | NR | NR | NR | NR | NR | NR | NR | NR | NR | NR | NR | NR | NR | NR | NR | NR | NR | NR | NR | NR | NR | NR | NR | NR | NR | NR | NR | NR | NR | NR | NR | NR |
| | | | | | Sym pt. | total | 5 | 2 | 3 | 5 | 70,0 | 60,9 | 5 | 233,4 | 320,7 | 5 | NR | NR | NR | 5 | 148,6 | 315,0 | 2,0 | 449,5 | 41,7 | 5 | 244,0 | 312,2 | 1 | 1 | 2,0 | NR | NR | 3 | NR | NR | NR | NR | NR | 2 | 5,0 | 1,4 | 2 | 68,5 | 44,5 |
| | | | | | Sym pt. | EO | 4 | 2 | 2 | 4 | 75,5 | 68,8 | 4 | 113,8 | 204,2 | 4 | NR | NR | NR | 4 | 7,8 | 4,6 | 2,0 | 449,5 | 41,7 | 4 | 127,0 | 196,5 | 1 | 1 | 2,0 | NR | NR | 2 | NR | NR | NR | NR | NR | 1 | 6,0 | NR | 1 | 100,0 | NR |
| | | | | | Sym pt. | LO | 1 | 0 | 1 | 1 | 48,0 | NR | 1 | 712,0 | NR | 1 | NR | NR | NR | 1 | 712,0 | NR | NR | NR | NR | 1 | 712,0 | NR | 0 | NR | NR | NR | NR | 1 | NR | NR | NR | NR | NR | 1 | 4,0 | NR | 1 | 37,0 | NR |
| Lee et al. 2007 | South Korea | 7 | 5 | 79,6 | NBS | total | 1 | 1 | 0 | 1 | 60,0 | NR | 1 | 7,0 | NR | 0 | 1 | 33,8 | NR | NR | NR | NR | NR | NR | NR | NR | NR | NR | 0 | NR | NR | NR | NR | 0 | NR | NR | NR | NR | NR | NR | NR | NR | NR | NR | NR |
| | | | | | NBS | EO | 0 | NR | NR | NR | NR | NR | NR | NR | NR | NR | NR | NR | NR | NR | NR | NR | NR | NR | NR | NR | NR | NR | NR | NR | NR | NR | NR | NR | NR | NR | NR | NR | NR | NR | NR | NR | NR | NR | NR | NR |
| | | | | | NBS | LO | 0 | NR | NR | NR | NR | NR | NR | NR | NR | NR | NR | NR | NR | NR | NR | NR | NR | NR | NR | NR | NR | NR | NR | NR | NR | NR | NR | NR | NR | NR | NR | NR | NR | NR | NR | NR | NR | NR | NR | NR |
| | | | | | NBS | asymptomatic | 1 | 1 | 0 | 1 | 60,0 | NR | 1 | 7,0 | NR | 0 | 1 | 33,8 | NR | NR | NR | NR | NR | NR | NR | NR | NR | NR | 0 | NR | NR | NR | NR | 0 | NR | NR | NR | NR | NR | NR | NR | NR | NR | NR | NR |

| | | | | | | | | | | | | | | | | | | | | | | | | | | | | | | | | | | | | | | | | | | | | | |
|---|---|---|---|---|---|---|---|---|---|---|---|---|---|---|---|---|---|---|---|---|---|---|---|---|---|---|---|---|---|---|---|---|---|---|---|---|---|---|---|---|---|---|---|---|---|
| | | | | | Sym pt. | total | 4 | 3 | 1 | 4 | 103,0 | 70,0 | 4 | 1340,5 | 1167,9 | 4 | 1 | 4,8 | NR | 1 | 10,0 | NR | NR | NR | NR | NR | NR | NR | 0 | NR | NR | NR | NR | 4 | NR | NR | NR | NR | NR | NR | NR | NR | NR | NR | NR |
| | | | | | Sym pt. | EO | 4 | 3 | 1 | 4 | 103,0 | 70,0 | 4 | 1340,5 | 1167,9 | 4 | 1 | 4,8 | NR | 1 | 10,0 | NR | NR | NR | NR | NR | NR | NR | 0 | NR | NR | NR | NR | 4 | NR | NR | NR | NR | NR | NR | NR | NR | NR | NR | NR |
| | | | | | Sym pt. | LO | 0 | NR | NR | NR | NR | NR | NR | NR | NR | NR | NR | NR | NR | NR | NR | NR | NR | NR | NR | NR | NR | NR | NR | NR | NR | NR | NR | NR | NR | NR | NR | NR | NR | NR | NR | NR | NR | NR | NR |
| Lin et al. 2007 | Taiwan | 6 | 6 | 68,7 | NBS | total | 4 | 1 | 3 | 4 | 46,5 | 30,3 | 4 | 26,8 | 24,4 | 0 | 1 | 7,4 | NR | NR | NR | NR | NR | NR | NR | 4 | 26,8 | 24,4 | 0 | NR | NR | NR | 0 | 0 | NR | NR | NR | NR | NR | NR | NR | NR | NR | NR | NR |
| | | | | | NBS | EO | 0 | NR | NR | NR | NR | NR | NR | NR | NR | NR | NR | NR | NR | NR | NR | NR | NR | NR | NR | NR | NR | NR | NR | NR | NR | NR | NR | NR | NR | NR | NR | NR | NR | NR | NR | NR | NR | NR | NR |
| | | | | | NBS | LO | 0 | NR | NR | NR | NR | NR | NR | NR | NR | NR | NR | NR | NR | NR | NR | NR | NR | NR | NR | NR | NR | NR | NR | NR | NR | NR | NR | NR | NR | NR | NR | NR | NR | NR | NR | NR | NR | NR | NR |
| | | | | | NBS | asymptomatic | 4 | 1 | 3 | 4 | 46,5 | 30,3 | 4 | 26,8 | 24,4 | 0 | 1 | 7,4 | NR | NR | NR | NR | NR | NR | NR | 4 | 26,8 | 24,4 | 0 | NR | NR | NR | 0 | 0 | NR | NR | NR | NR | NR | NR | NR | NR | NR | NR | NR |
| | | | | | Sym pt. | total | 2 | 1 | 1 | 2 | 258,0 | 8,5 | 2 | 890,0 | 251,7 | 2 | NR | NR | NR | NR | NR | NR | NR | NR | NR | 2 | 890,0 | 251,7 | 0 | NR | NR | NR | 2 | 2 | NR | NR | NR | NR | NR | NR | NR | NR | NR | NR | NR |
| | | | | | Sym pt. | EO | 0 | NR | NR | NR | NR | NR | NR | NR | NR | NR | NR | NR | NR | NR | NR | NR | NR | NR | NR | NR | NR | NR | NR | NR | NR | NR | NR | NR | NR | NR | NR | NR | NR | NR | NR | NR | NR | NR | NR |
| | | | | | Sym pt. | LO | 2 | 1 | 1 | 2 | 258,0 | 8,5 | 2 | 890,0 | 251,7 | 2 | NR | NR | NR | NR | NR | NR | NR | NR | NR | 2 | 890,0 | 251,7 | 0 | NR | NR | NR | 2 | 2 | NR | NR | NR | NR | NR | NR | NR | NR | NR | NR | NR |
| Tokatli et al. 1998 | Turkey | 6 | 6 | 56,3 | NBS | total | 0 | NR | NR | NR | NR | NR | NR | NR | NR | NR | NR | NR | NR | NR | NR | NR | NR | NR | NR | NR | NR | NR | NR | NR | NR | NR | NR | NR | NR | NR | NR | NR | NR | NR | NR | NR | NR | NR | NR |
| | | | | | NBS | EO | 0 | NR | NR | NR | NR | NR | NR | NR | NR | NR | NR | NR | NR | NR | NR | NR | NR | NR | NR | NR | NR | NR | NR | NR | NR | NR | NR | NR | NR | NR | NR | NR | NR | NR | NR | NR | NR | NR | NR |
| | | | | | NBS | LO | 0 | NR | NR | NR | NR | NR | NR | NR | NR | NR | NR | NR | NR | NR | NR | NR | NR | NR | NR | NR | NR | NR | NR | NR | NR | NR | NR | NR | NR | NR | NR | NR | NR | NR | NR | NR | NR | NR | NR |
| | | | | | NBS | asymptomatic | 0 | NR | NR | NR | NR | NR | NR | NR | NR | NR | NR | NR | NR | NR | NR | NR | NR | NR | NR | NR | NR | NR | NR | NR | NR | NR | NR | NR | NR | NR | NR | NR | NR | NR | NR | NR | NR | NR | NR |
| | | | | | Sym pt. | total | 6 | 4 | 2 | 6 | 63,2 | 61,9 | 4 | 1165,5 | 676,3 | 6 | NR | NR | NR | 6 | 477,3 | 570,5 | NR | NR | NR | 3 | 1257,3 | 797,2 | 3 | 3 | 10,3 | 17,0 | 1 | 1 | NR | NR | NR | NR | NR | NR | NR | NR | NR | NR | NR |
| | | | | | Sym pt. | EO | 2 | 1 | 1 | 2 | 0,5 | 0,4 | NR | NR | NR | 2 | NR | NR | NR | 2 | 5,0 | 2,8 | NR | NR | NR | NA | NA | NA | 2 | 2 | 0,5 | 0,4 | 0 | 0 | NR | NR | NR | NR | NR | NR | NR | NR | NR | NR | NR |
| | | | | | Sym pt. | LO | 4 | 3 | 1 | 4 | 94,5 | 49,6 | 4 | 1165,5 | 676,3 | 4 | NR | NR | NR | 4 | 713,5 | 565,1 | NR | NR | NR | 3 | 1257,3 | 797,2 | 1 | 1 | 30,0 | NA | 1 | 1 | NR | NR | NR | NR | NR | NR | NR | NR | NR | NR | NR |
| Dodelson de Kremer et al. 1992 | Argentina | 5 | 5 | 57,4 | NBS | total | 0 | NR | NR | NR | NR | NR | NR | NR | NR | NR | NR | NR | NR | NR | NR | NR | NR | NR | NR | NR | NR | NR | NR | NR | NR | NR | NR | NR | NR | NR | NR | NR | NR | NR | NR | NR | NR | NR | NR |
| | | | | | NBS | EO | 0 | NR | NR | NR | NR | NR | NR | NR | NR | NR | NR | NR | NR | NR | NR | NR | NR | NR | NR | NR | NR | NR | NR | NR | NR | NR | NR | NR | NR | NR | NR | NR | NR | NR | NR | NR | NR | NR | NR |
| | | | | | NBS | LO | 0 | NR | NR | NR | NR | NR | NR | NR | NR | NR | NR | NR | NR | NR | NR | NR | NR | NR | NR | NR | NR | NR | NR | NR | NR | NR | NR | NR | NR | NR | NR | NR | NR | NR | NR | NR | NR | NR | NR |
| | | | | | NBS | asymptomatic | 0 | NR | NR | NR | NR | NR | NR | NR | NR | NR | NR | NR | NR | NR | NR | NR | NR | NR | NR | NR | NR | NR | NR | NR | NR | NR | NR | NR | NR | NR | NR | NR | NR | NR | NR | NR | NR | NR | NR |
| | | | | | Sym pt. | total | 5 | 2 | 3 | 5 | 115,2 | 69,2 | 5 | 482,2 | 644,7 | 5 | NR | NR | NR | 5 | 637,6 | 412,3 | 1 | 129,0 | NR | 5 | 482,2 | 644,7 | 1 | 1 | 36,0 | NR | 3 | 3 | NR | NR | NR | NR | NR | 2 | 9,2 | 1,7 | 2 | 65,5 | 12,0 |
| | | | | | Sym pt. | EO | 1 | 1 | 0 | 1 | 72,0 | NR | 1 | 12,0 | NR | 1 | NR | NR | NR | 1 | 8,0 | NA | 1 | 129,0 | NR | 1 | 12,0 | NA | 0 | NA | NA | NR | 1 | 1 | NR | NR | NR | NR | NR | NR | NR | NR | NR | NR | NR |
| | | | | | Sym pt. | LO | 4 | 1 | 3 | 4 | 126,0 | 74,9 | 4 | 600,0 | 679,7 | 4 | NR | NR | NR | 4 | 795,0 | 248,0 | NA | NR | NR | 4 | 600,0 | 679,7 | 1 | 1 | 36,0 | NR | 2 | 2 | NR | NR | NR | NR | NR | 2 | 9,2 | 1,7 | 2 | 65,5 | 12,0 |
| Tanaka 1990 | USA | 37 | 37 | 73,7 | NBS | total | 0 | NR | NR | NR | NR | NR | NR | NR | NR | NR | NR | NR | NR | NR | NR | NR | NR | NR | NR | NR | NR | NR | NR | NR | NR | NR | NR | NR | NR | NR | NR | NR | NR | NR | NR | NR | NR | NR | NR |
| | | | | | NBS | EO | 0 | NR | NR | NR | NR | NR | NR | NR | NR | NR | NR | NR | NR | NR | NR | NR | NR | NR | NR | NR | NR | NR | NR | NR | NR | NR | NR | NR | NR | NR | NR | NR | NR | NR | NR | NR | NR | NR | NR |
| | | | | | NBS | LO | 0 | NR | NR | NR | NR | NR | NR | NR | NR | NR | NR | NR | NR | NR | NR | NR | NR | NR | NR | NR | NR | NR | NR | NR | NR | NR | NR | NR | NR | NR | NR | NR | NR | NR | NR | NR | NR | NR | NR |
| | | | | | NBS | asymptomatic | 0 | NR | NR | NR | NR | NR | NR | NR | NR | NR | NR | NR | NR | NR | NR | NR | NR | NR | NR | NR | NR | NR | NR | NR | NR | NR | NR | NR | NR | NR | NR | NR | NR | NR | NR | NR | NR | NR | NR |
| | | | | | Sym pt. | total | 37 | 18 | 19 | NR | NR | NR | NR | NR | NR | 37 | NR | NR | NR | NR | NR | NR | NR | NR | NR | NR | NR | NR | 16 | NR | NR | NR | NR | NR | NR | NR | NR | NR | NR | NR | NR | NR | NR | NR | NR |

| | | | | | | | | | | | | | | | | | | | | | | | | | | | | | | | | | | | | | | | | | | | | | |
|---|---|---|---|---|---|---|---|---|---|---|---|---|---|---|---|---|---|---|---|---|---|---|---|---|---|---|---|---|---|---|---|---|---|---|---|---|---|---|---|---|---|---|---|---|---|
| | | | | | Sympt. | EO | 28 | NR | NR | NR | NR | NR | NR | NR | NR | 28 | NR | NR | NR | NR | NR | NR | NR | NR | NR | NR | NR | NR | NR | NR | NR | NR | NR | NR | NR | NR | NR | NR | NR | NR | NR | NR | NR | NR | NR |
| | | | | | Sympt. | LO | 9 | NR | NR | NR | NR | NR | NR | NR | NR | 9 | NR | NR | NR | NR | NR | NR | NR | NR | NR | NR | NR | NR | NR | NR | NR | NR | NR | NR | NR | NR | NR | NR | NR | NR | NR | NR | NR | NR | NR |
| Berry et al. 1988 | USA | 9 | 9 | 73,7 | NBS | total | 0 | NR | NR | NR | NR | NR | NR | NR | NR | NR | NR | NR | NR | NR | NR | NR | NR | NR | NR | NR | NR | NR | NR | NR | NR | NR | NR | NR | NR | NR | NR | NR | NR | NR | NR | NR | NR | NR | NR | NR | NR |
| | | | | | NBS | EO | 0 | NR | NR | NR | NR | NR | NR | NR | NR | NR | NR | NR | NR | NR | NR | NR | NR | NR | NR | NR | NR | NR | NR | NR | NR | NR | NR | NR | NR | NR | NR | NR | NR | NR | NR | NR | NR | NR | NR | NR | NR |
| | | | | | NBS | LO | 0 | NR | NR | NR | NR | NR | NR | NR | NR | NR | NR | NR | NR | NR | NR | NR | NR | NR | NR | NR | NR | NR | NR | NR | NR | NR | NR | NR | NR | NR | NR | NR | NR | NR | NR | NR | NR | NR | NR | NR | NR |
| | | | | | NBS | asymptomatic | 0 | NR | NR | NR | NR | NR | NR | NR | NR | NR | NR | NR | NR | NR | NR | NR | NR | NR | NR | NR | NR | NR | NR | NR | NR | NR | NR | NR | NR | NR | NR | NR | NR | NR | NR | NR | NR | NR | NR | NR | NR |
| | | | | | Sympt. | total | 9 | 5 | 4 | 9 | 94,0 | 52,0 | 9 | 392,3 | 578,2 | 8 | NR | NR | NR | 5 | 4,6 | 4,8 | NR | NR | NR | 9 | 392,3 | 578,2 | 0 | NR | NR | NR | 6 | 6 | NR | NR | NR | NR | NR | 9 | 9,0 | 6,2 | 9 | 80,6 | 21,1 |
| | | | | | Sympt. | EO | 5 | 4 | 1 | 4 | 70,4 | 54,4 | 5 | 10,2 | 5,5 | 5 | NR | NR | NR | 5 | 4,6 | 4,8 | NR | NR | NR | 5 | 10,2 | 5,5 | 0 | NR | NR | NR | 3 | 3 | NR | NR | NR | NR | NR | 5 | 4,3 | 3,6 | 5 | 83,8 | 26,2 |
| | | | | | Sympt. | LO | 4 | 1 | 3 | 4 | 123,5 | 34,3 | 4 | 870,0 | 586,3 | 3 | NR | NR | NR | NR | NR | NR | NR | NR | NR | 4 | 870,0 | 586,3 | 0 | NR | NR | NR | 3 | 3 | NR | NR | NR | NR | NR | 4 | 8,7 | 2,7 | 4 | 76,5 | 15,4 |

**Supplementary Table 3: Suggested core outcome set for classic IVA** containing elements from six core areas, along with definitions and guidance on measurement and implementation. This is inspired by the core outcome sets for medium-chain Acyl-CoA dehydrogenase deficiency and phenylketonuria (3-5) and was created based on the suggested core outcome set for maple syrup urine disease (6).

| Core Area | Outcome | Definition | Suggested implementation |
|---|---|---|---|
| **Growth and development** | Overall child development and functioning | Process of change across a combination of physical, emotional, cognitive, social, and language areas over the course of childhood, including ability to perform age-appropriate daily activities and functions | i.e. Vineland Adaptive Behaviour Scales (7), Denver Developmental Screening Test (8) |
| | Cognition and intelligence | A child's learning abilities related to the activity and development of the brain | Age-adapted IQ-based psychological test (i.e. Wechsler Preschool and Primary Scale of Intelligence (WPPSI) (9), Wechsler Adult Intelligence Scale (WAIS) (10, 11), Wechsler Intelligence Scale for Children (WISC) (12, 13), Bayley Scales of Infant Development (BSID) (14), Denver Developmental Screening Test (8) |
| **Life impact** | Child quality of life | A general concept of well-being that refers to a combination of various life aspects (e.g. health, emotional, and social), is assessed from the child's perspective | Pediatric Quality of Life Inventory (PedsQL) (15) |
| | Disease-specific child quality of life | A disease-specific concept of well-being that refers to a combination of various life aspects (e.g., physical, mental, and social), refers to intoxication-type inherited metabolic diseases, is assessed from the child's perspective | Disease-specific Health-related Quality of Life Questionnaire for Intoxication-type Inborn Errors of Metabolism (MetabQoL 1.0) (16) |
| **Pathophysiological manifestations** | Metabolic decompensation | A serious health condition caused by the build-up of toxic substances in the blood, often triggered by catabolic states like fasting, (febrile) illness or increased protein intake in people with classic IVA (17) | Presence of metabolic acidosis and hyperammonemia, and/or encephalopathy (vomiting, altered mental status, reduced consciousness or coma, lethargy) and a distinct odor of sweaty feet) (17, 18); administrative data and/or parent/patient report on frequency and severity |
| | Isovaleryl-carnitine in dried blood spots (DBS) and isovaleryl-glycine in urine | Concentration of isovaleryl-carnitine in DBS and isovaleryl-glycine in urine and whether the level is consistent with the targeted level (19) | Report in SI-units |

| | | | |
|---|---|---|---|
| | Disease-specific clinical findings | Permanent or transient symptoms possibly related to classic IVA | Administrative data, physical examination; HPO-coded |
| **Resource use** | Emergency department use and hospitalisations | Measures related to the frequency and nature of visits to the emergency department and hospitalisations | Administrative data and/or parent/patient report |
| | Access to care | The extent or ease with which patients or communities can use appropriate health services in proportion to their needs, with a specific focus on health system capacity and the resources required to deliver care | The operational definition is likely to be context and/or study-dependent, i.e. Health access and quality index (HAQ) according to country (20-22) |
| **Long-term management** | Adherence to classic IVA treatment plan | The extent to which someone sticks to a special diet (e.g. leucin or protein reduced diet, leucine-free amino acid supplementation, detoxification by carnitine or glycine supplementation the avoidance of catabolic episodes) (18, 23) or other aspects of the treatment plan (e.g. regular blood/urine samples) prescribed specifically for someone with classic IVA | Actual vs. required number of blood/urine samples, actual vs. prescribed protein intake (24-hours-recall or 3-days-food diary), actual vs. prescribed intake of leucine-free amino acid mixture (self-/parent-reported compliance) |
| **Mortality** | Death | The end of a person’s life | Administrative data on age at death and cause |
